# Eleven Years of Change: Disease Progression in Biomarker-Defined Sporadic Parkinson’s Disease

**DOI:** 10.1101/2024.10.09.24315191

**Authors:** Paulina Gonzalez-Latapi, Caroline Gochanour, Hyunkeun Cho, Seung Ho Choi, Chelsea Caspell-Garcia, Christopher Coffey, Michael Brumm, David-Erick Lafontant, Yuge Xiao, Caroline Tanner, Charles S. Venuto, Karl Kieburtz, Lana M. Chahine, Kathleen L Poston, Andrew Siderowf, Ken Marek, Tanya Simuni, Parkinson’s Progression Markers Initiative

## Abstract

Long-term longitudinal data on outcomes in sporadic Parkinson’s Disease are limited, especially from cohorts with extensive biological characterization. Recent advances in biomarkers characterization of Parkinson’s Disease necessitate an updated examination of long-term progression within contemporary cohorts like the Parkinson’s Progression Markers Initiative, which enrolled individuals within 2 years of clinical diagnosis of Parkinson’s Disease. Our study leverages the Neuronal Synuclein Disease framework, which defines the disease based on biomarker assessed presence of neuronal alpha-synuclein and dopamine deficit, rather than based on conventional clinical diagnostic criteria. In this study we aimed to provide a comprehensive long-term description of disease progression using the integrated biological and clinical staging system framework.

We analyzed data from 344 participants from the sporadic Parkinson’s Disease cohort in the Parkinson’s Progression Markers Initiative, who met Neuronal Synuclein Disease criteria. We assessed 11-year progression in a spectrum of clinical measures. We used Cox proportional hazards models to assess the association between baseline stage and time to key outcomes, including survival, postural instability (Hoehn & Yahr ≥ 3), loss of independence (Schwab & England < 80%), cognitive decline, and domain-based milestones such as walking and balance, motor complications, autonomic dysfunction, and activities of daily living. Additional analyses were completed to account for death and participant dropout. Biomarker analysis included dopamine transporter binding measures, as well as serum urate, neurofilament light chain and CSF amyloid-beta, phosphorylated tau and total tau.

At baseline, despite the cohort consisting of individuals within 2 years of clinical diagnosis, there was clear separation of participants in Neuronal Synuclein Disease Stages (23% Stage 2b, 67% Stage 3, 10% Stage 4). At 11 years, data were available for 153 participants; 35 participants had died over the follow up period. Of retained participants, 59% presented normal cognition, 24% had evidence of postural instability and mean Schwab & England score was 78.5. Serum neurofilament light chain consistently increased over time. No other biofluids had a consistent change in trajectory. Of importance, baseline Neuronal Synuclein Disease Stage predicted progression to clinically meaningful milestones.

This study provides data on longitudinal, 11-year progression in Neuronal Synuclein Disease participants within 2 years of clinical diagnosis. We observed better long-term outcomes in this contemporary observational study cohort. It highlights the heterogeneity in the early Parkinson’s Disease population as defined by clinical diagnostic criteria and underscores the importance of shifting from clinical to biologically and functionally based inclusion criteria in the design of new clinical trials.

## Introduction

Parkinson’s Disease has been historically defined by clinical features, with a definite diagnosis only possible through the identification of alpha-synuclein (α-syn) in Lewy bodies and nigrostriatal degeneration in brain autopsy.^1–3^ Clinical diagnostic criteria are based solely on the presence of motor features,^4^ which are only apparent once around 70-80% of dopaminergic neurons are lost.^5^ Thus, diagnosis is made only once the disease process has been ongoing for more than a decade.^6^ Additionally, there is substantial heterogeneity in the clinical presentation and rates of progression among Parkinson’s Disease patients.^7^

Despite a significant number of observational and interventional studies, long term data on Parkinson’s Disease progression remains scarce. One of the most referenced cohorts, the Sydney Multicenter Study,^8–10^ did not include biological characterization. In addition, while follow-up spanned 20 years, only a small proportion of the original cohort remained by the end of the study. Moreover, since the publication of the Sydney data, our understanding of Parkinson’s Disease and its management has evolved substantially. For instance, Gallagher *et al*^11^ report findings from two large prospective Parkinson’s Disease cohorts, showing a lower frequency of dementia and a longer progression period before its onset than previously suggested by older studies. Thus, a description of the disease within a contemporary cohort, followed longitudinally with clinical and biological data, may inform an evolving perspective on the course of Parkinson’s Disease under contemporary management paradigms.

Recently, the detection of neuronal α-syn through seed amplification assay in CSF (CSF-SAA) has emerged as a reliable in vivo biomarker of synuclein pathology, validated in multiple cohorts^12–15^ and against postmortem tissue.^16,17^ Based on these findings, the concept of neuronal synuclein disease (NSD) was proposed, defined by the presence of neuronal α-syn as measured by a validated biomarker, such as CSF-SAA and stage-dependent evidence of dopaminergic dysfunction, as measured by a validated biomarker such as dopamine transporter (DAT binding) deficit. Simuni *et al*^18^ have proposed an integrated biological and clinical staging system for NSD (NSD-ISS), which builds on similar efforts in other neurodegenerative diseases, including Alzheimer’s disease^19,20^ and Huntington’s disease.^21^ The NSD-ISS includes 7 stages, defined by each biomarker and presence of clinical features and their functional consequences.

The Parkinson’s Progression Markers Initiative (PPMI) provides a uniquely comprehensive set of longitudinal, clinical, imaging and biosample data from participants recruited as clinically defined de novo Parkinson’s Disease, at-risk individuals and healthy controls.^22^ PPMI has followed participants prospectively for around 13 years since its inception. Among the 373 individuals who were enrolled within 2 years of a diagnosis of Parkinson’s Disease based on clinical criteria, who were sporadic (without identified pathogenic variants) and with enough data to determine NSD staging, 93% meet NSD criteria.^18^ By analyzing PPMI participants who meet NSD criteria, we are able to conduct a detailed, long-term longitudinal study within a biologically homogeneous population.

The aims of our study were to: (i) describe long term outcomes of NSD participants originally enrolled in the sporadic Parkinson’s Disease cohort in PPMI; (ii) analyze the association between baseline NSD stage and survival; (iii) analyze the association between baseline NSD stage and key outcomes which have the greatest impact on participants’ quality of life and function, namely: development of postural instability, loss of independence and cognitive decline; and (iv) to analyze the association between baseline NSD stage and the time to reach domain-based disease milestones as described by Brumm et al.^23^

## Materials and methods

### PPMI

PPMI is an ongoing international, multicenter, prospective cohort study initiated in June 2010 as described previously.^22,24^ The study was approved by the institutional review board at each site, and all participants provided written informed consent. The primary aim of PPMI is to identify genomic, biochemical, or imaging biomarkers of clinical progression. The detailed study protocol, manuals, biofluid collection, and storage processes are available at www.ppmi-info.org/study-design.

### Study population

Participants included in the study were those enrolled in the original sporadic Parkinson’s Disease cohort in PPMI. Briefly, enrollment criteria to the sporadic Parkinson’s Disease cohort included: (i) presence of two or more of the following: bradykinesia, rigidity, and resting tremor OR presence of either an asymmetric resting tremor or asymmetric bradykinesia (ii) disease duration from diagnosis of ≤2 years, (iii) DAT binding deficit based visual interpretation. Participants could not be treated with dopaminergic therapy or expected to need treatment within 6 months of enrollment. From this group, we selected individuals that fulfilled NSD criteria at time of or within 12 months of enrollment into PPMI, and who were recruited prior to 2020 to allow for at least 5 years of longitudinal data.

### NSD-ISS staging

NSD is defined by pathologic neuronal α-syn (S) and eventual dopaminergic neuronal dysfunction (D), independent of clinical features. The NSD-ISS integrates these biological anchors and the degree of functional impairment as follows: Stage 2A (S+, D-, subtle signs/symptoms, no functional impairment), Stage 2B (S+, D+, subtle signs/symptoms, no functional impairment), Stage 3 (S+, D+, signs/symptoms +, slight functional impairment), Stage 4 or 5 (S+, D+, signs/symptoms +, mild or moderate functional impairment) **(Supplementary Table 1).** If participants completed a follow-up visit but were missing one or more of the components used to determine stage, the data needed to assign a stage was carried forward from the previous visit.

### Clinical Assessments

PPMI includes a wide array of investigator completed and participant reported measures of motor, non-motor and cognitive function. For this analysis we utilized demographic data (age, sex, race, time since diagnosis, education level), Movement Disorder Society-Unified Parkinson’s Disease Rating Scale (MDS-UPDRS)-Parts I-IV,^25^ Hoehn & Yahr (H&Y),^26^ Schwab-England activities of daily living score (S&E),^27^ age/sex adjusted University of Pennsylvania Smell Identification Test (UPSIT) score,^28^ Scales for Outcomes in Parkinson’s disease-Autonomic (SCOPA-AUT) total score,^29^ Geriatric Depression Scale (GDS) score,^30^ REM sleep behavior disorder (RBD) screening questionnaire (RBDSQ) score,^31^ Levodopa Equivalent Daily Dose (LEDD).^32^ Cognition was measured with the Montreal Cognitive Assessment (MoCA)^33^ scores and site investigator’s clinical diagnosis of cognitive state (normal cognition, mild cognitive impairment [MCI] or Parkinson’s Disease dementia [PDD]), which was not fully implemented until study year 3.^34^ The site investigator is provided a guidance document on how to assess for subjective cognitive change compared with pre-Parkinson’s Disease state, impairment in cognitive abilities, and functional impairment due to cognitive deficits (i.e., providing specific examples of how cognitive impairment might adversely impact instrumental activities of daily living requiring cognitive abilities), with the option to review cognitive test results (e.g., MoCA, Hopkins Verbal Learning Test-Revised [HVLT-R],^35^ Benton Judgment of Line Orientation [JLO],^36^ Symbol-Digit Modalities Test [SDMT],^37^ Letter-Number Sequencing [LNS],^38^ and category [animal] fluency^39^). The guidance document was meant to approximate Parkinson’s Disease-MCI^40^ or Parkinson’s Disease-PDD^41^ criteria. All assessments were conducted annually.

We also analyzed time to reach domain-based disease milestones as described by Brumm *et al*.^23^ Briefly, these include 25 progression milestones, spanning six clinical domains: “walking and balance”; “motor complications”; “cognition”; “autonomic dysfunction”; “functional dependence”; and “activities of daily living” **(Supplementary Table 2).**

### Imaging biomarkers

Degree of dopaminergic dysfunction was assessed with DAT binding and quantified in two ways; as the striatal binding ratio (SBR) obtained from the ipsilateral putamen alone and as the average of SBRs obtained from the caudate and putamen in both hemispheres. DAT binding was assessed at baseline and at years 1, 2, and 4 of the study. Imaging acquisition and analysis protocols can be found at ppmi-info.org.

### Biofluid biomarkers

CSF amyloid-beta 1-42 (Aβ 1–42), total-tau, tau phosphorylated at threonine 181 position (p-tau) were measured at baseline and annually until year 9. Serum neurofilament light chain (NfL) was measured at baseline and then at years 1, 2, 3 and 5. Cutoffs for CSF Aβ 1–42 ≤ 683 pg/mL, p-tau ≥ 13 pg/mL and serum NfL ≥ 19.05 pg/mL were also used as variables for the analysis.^42^ Serum urate was measured at baseline and then annually until year 10. Due to the high degree of missingness, only measures up to year 5 were included in this analysis.

### Survival endpoints

The following endpoints were used to capture disease progression in our analyses: Death, postural instability (defined by H&Y ≥ 3), disability (defined as S&E < 80%) and cognitive decline (defined as site investigator’s clinical diagnosis of MCI or PDD). We analyzed time to reach five of the progression milestone domains as described by Brumm et al,^23^ including “walking and balance”; “motor complications”; “cognition”; “autonomic dysfunction”, and “activities of daily living”.

To address concerns about informative dropout, participants who withdrew were categorized by reason for withdrawal into non-informative and informative categories. Participants who withdrew due to operational reasons (completed study per-protocol; family, care-partner, or social issues; non-compliance with study procedures; transportation/travel issues; investigator or informant/caregiver decision; burden of study procedures) were considered non-informative or missing at random. Participants who withdrew due to adverse events, death, decline in health, withdrawal of consent, loss to follow-up, or other unspecified reasons were conservatively categorized as informative or missing not at random.

### Statistical analysis

Baseline demographic, clinical, and biological characteristics were reported for all participants and separately by baseline NSD stage. Frequency and percent were reported for categorical measures and median and range for continuous measures. To evaluate differences in baseline characteristics by NSD stage, Chi-Square and Fisher’s exact test (when appropriate) were presented for categorical measures and Kruskal-Wallis tests for continuous variables. Nonparametric tests were calculated for continuous measures due to the small number of participants in the Stage 4 group. Longitudinal measures were reported for each annual study visit up to year 11, which is the minimum annual visit for which all enrolled participants were eligible.

To evaluate participant progression over time by baseline stage, we calculated time from enrollment in PPMI to reaching the nine endpoints of interest: death, reaching H&Y stage ≥ 3, S&E <80%, cognitive decline, walking and balance (domain 1), motor complications (domain 2), cognition (domain 3), autonomic dysfunction (domain 4), and activities of daily living (domain 6). Cox proportional hazards models were used to model time from enrollment to each outcome, stratified by baseline stage, and adjusting for baseline age, sex, and education (<12 years or ≥12 years). If participants did not have an event during the follow-up period, they were censored at the time of their last assessment. If participants reached any of the nine endpoints at the baseline visit, they were removed from all models. Hazards ratios for comparing stage 2b to 3 and 2b to 4 are presented with their 95% Wald confidence intervals. Age, sex, and education adjusted survival curves are presented. We use a Bonferroni adjusted α-level of 0.0056 to adjust for multiple comparisons in survival analyses.

To assess the impact of death and informative withdrawal on our estimates of participant progression, we conducted a sensitivity analysis using Fine and Gray subdistributional hazards models for time from enrollment to each of the nine endpoints of interest, with death and informative withdrawal as competing outcomes (for the survival outcome, informative withdrawal was the only competing event). Models included the same population and covariates as in the Cox regression analysis. Subdistributional hazards ratios for comparing stage 2b to 3 and 2b to 4 are presented with their 95% Wald confidence intervals.

All analyses were performed using SAS 9.4 (SAS/STAT 15.3; SAS Institute Inc., Cary, NC).

## Results

### Baseline Demographic Characteristics of NSD participants

PPMI enrolled *n*=423 sporadic Parkinson’s Disease participants prior to 2020, *n*=79 were excluded from this analysis (*n=*33 carried a genetic variant associated with Parkinson’s Disease, *n=*17 did not have enough data to determine NSD staging, *n=*26 did not fulfill NSD criteria, *n=*1 removed due to a data error and only *n=*2 fulfilled criteria for NSD Stage 2a and were not included in the analysis). The final analytic sample included a total of *n=*344 NSD participants, *n=*225 (65%) male, with a median [min-max] age of 62.4 [33.7 – 84.9] years at time of enrollment and a time from diagnosis of 0.3 years [0-3 years]. There were no demographic differences at baseline between different NSD stages **(Table 1)**.

**Table 1.**
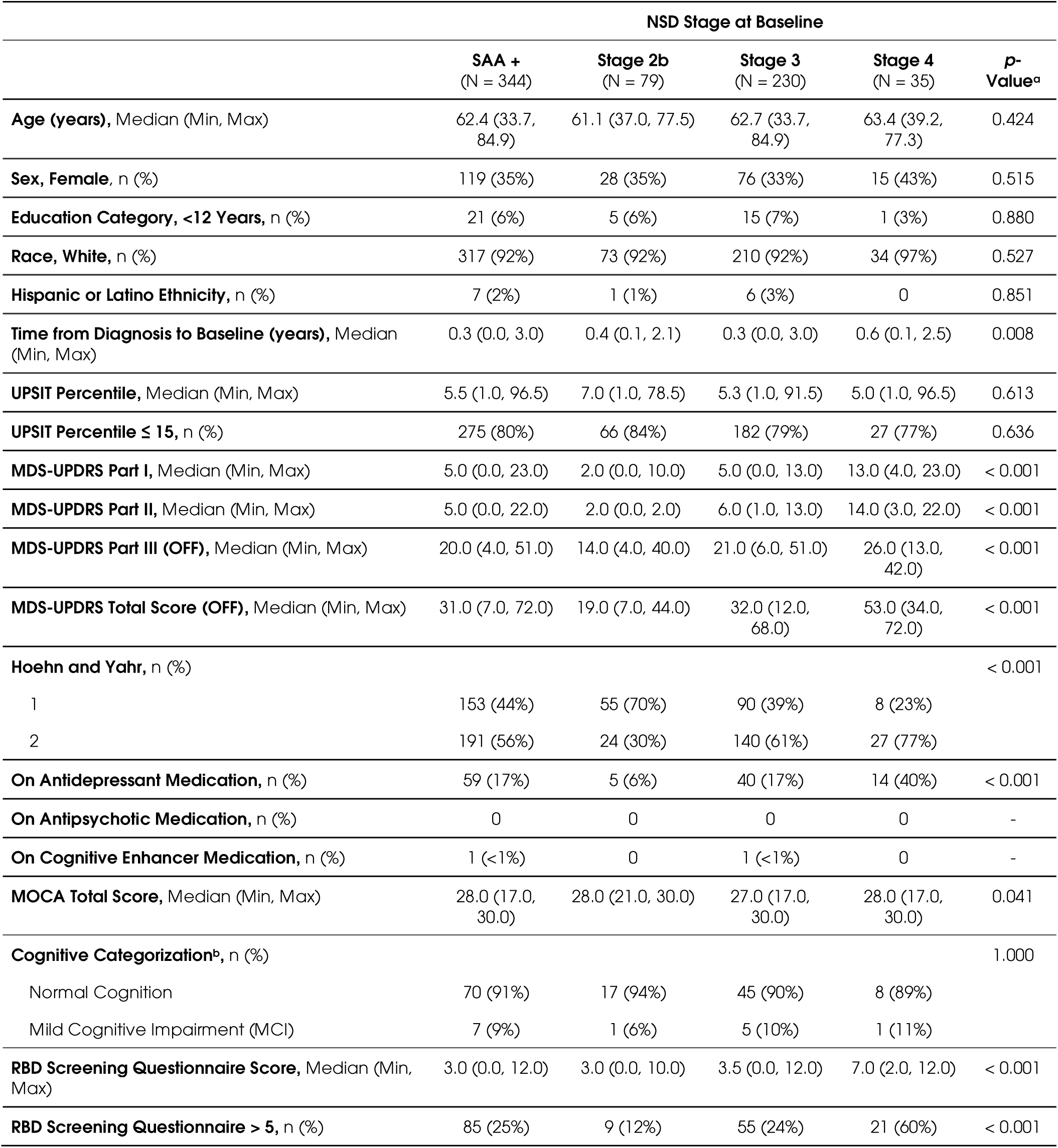
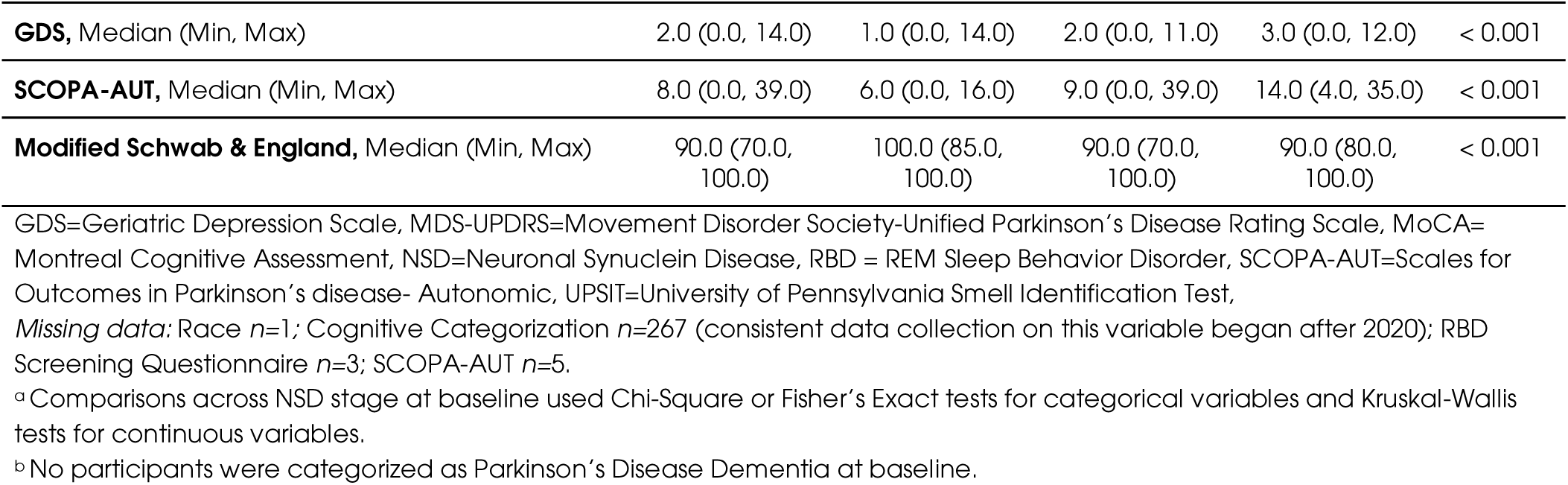
Baseline Demographic and Clinical Characteristics.

|  | NSD Stage at Baseline |  |  |  |  |
| --- | --- | --- | --- | --- | --- |
|  | SAA +<br>(N = 344) | Stage 2b<br>(N = 79) | Stage 3<br>(N = 230) | Stage 4<br>(N = 35) | p-Value<sup>a</sup> |
| **Age (years)**, Median (Min, Max) | 62.4 (33.7, 84.9) | 61.1 (37.0, 77.5) | 62.7 (33.7, 84.9) | 63.4 (39.2, 77.3) | 0.424 |
| **Sex, Female**, n (%) | 119 (35%) | 28 (35%) | 76 (33%) | 15 (43%) | 0.515 |
| **Education Category, <12 Years**, n (%) | 21 (6%) | 5 (6%) | 15 (7%) | 1 (3%) | 0.880 |
| **Race, White**, n (%) | 317 (92%) | 73 (92%) | 210 (92%) | 34 (97%) | 0.527 |
| **Hispanic or Latino Ethnicity**, n (%) | 7 (2%) | 1 (1%) | 6 (3%) | 0 | 0.851 |
| **Time from Diagnosis to Baseline (years)**, Median (Min, Max) | 0.3 (0.0, 3.0) | 0.4 (0.1, 2.1) | 0.3 (0.0, 3.0) | 0.6 (0.1, 2.5) | 0.008 |
| **UPSIT Percentile**, Median (Min, Max) | 5.5 (1.0, 96.5) | 7.0 (1.0, 78.5) | 5.3 (1.0, 91.5) | 5.0 (1.0, 96.5) | 0.613 |
| **UPSIT Percentile ≤ 15**, n (%) | 275 (80%) | 66 (84%) | 182 (79%) | 27 (77%) | 0.636 |
| **MDS-UPDRS Part I**, Median (Min, Max) | 5.0 (0.0, 23.0) | 2.0 (0.0, 10.0) | 5.0 (0.0, 13.0) | 13.0 (4.0, 23.0) | < 0.001 |
| **MDS-UPDRS Part II**, Median (Min, Max) | 5.0 (0.0, 22.0) | 2.0 (0.0, 2.0) | 6.0 (1.0, 13.0) | 14.0 (3.0, 22.0) | < 0.001 |
| **MDS-UPDRS Part III (OFF)**, Median (Min, Max) | 20.0 (4.0, 51.0) | 14.0 (4.0, 40.0) | 21.0 (6.0, 51.0) | 26.0 (13.0, 42.0) | < 0.001 |
| **MDS-UPDRS Total Score (OFF)**, Median (Min, Max) | 31.0 (7.0, 72.0) | 19.0 (7.0, 44.0) | 32.0 (12.0, 68.0) | 53.0 (34.0, 72.0) | < 0.001 |
| **Hoehn and Yahr**, n (%) |  |  |  |  | < 0.001 |
| 1 | 153 (44%) | 55 (70%) | 90 (39%) | 8 (23%) |  |
| 2 | 191 (56%) | 24 (30%) | 140 (61%) | 27 (77%) |  |
| **On Antidepressant Medication**, n (%) | 59 (17%) | 5 (6%) | 40 (17%) | 14 (40%) | < 0.001 |
| **On Antipsychotic Medication**, n (%) | 0 | 0 | 0 | 0 | - |
| **On Cognitive Enhancer Medication**, n (%) | 1 (<1%) | 0 | 1 (<1%) | 0 | - |
| **MOCA Total Score**, Median (Min, Max) | 28.0 (17.0, 30.0) | 28.0 (21.0, 30.0) | 27.0 (17.0, 30.0) | 28.0 (17.0, 30.0) | 0.041 |
| **Cognitive Categorization<sup>b</sup>**, n (%) |  |  |  |  | 1.000 |
| Normal Cognition | 70 (91%) | 17 (94%) | 45 (90%) | 8 (89%) |  |
| Mild Cognitive Impairment (MCI) | 7 (9%) | 1 (6%) | 5 (10%) | 1 (11%) |  |
| **RBD Screening Questionnaire Score**, Median (Min, Max) | 3.0 (0.0, 12.0) | 3.0 (0.0, 10.0) | 3.5 (0.0, 12.0) | 7.0 (2.0, 12.0) | < 0.001 |
| **RBD Screening Questionnaire > 5**, n (%) | 85 (25%) | 9 (12%) | 55 (24%) | 21 (60%) | < 0.001 |
| **GDS**, Median (Min, Max) | 2.0 (0.0, 14.0) | 1.0 (0.0, 14.0) | 2.0 (0.0, 11.0) | 3.0 (0.0, 12.0) | < 0.001 |
| **SCOPA-AUT**, Median (Min, Max) | 8.0 (0.0, 39.0) | 6.0 (0.0, 16.0) | 9.0 (0.0, 39.0) | 14.0 (4.0, 35.0) | < 0.001 |
| **Modified Schwab & England**, Median (Min, Max) | 90.0 (70.0, 100.0) | 100.0 (85.0, 100.0) | 90.0 (70.0, 100.0) | 90.0 (80.0, 100.0) | < 0.001 |

GDS=Geriatric Depression Scale, MDS-UPDRS=Movement Disorder Society-Unified Parkinson's Disease Rating Scale, MoCA=Montreal Cognitive Assessment, NSD=Neuronal Synuclein Disease, RBD = REM Sleep Behavior Disorder, SCOPA-AUT=Scales for Outcomes in Parkinson's disease- Autonomic, UPSIT=University of Pennsylvania Smell Identification Test,
*Missing data:* Race $n=1$ ; Cognitive Categorization $n=267$ (consistent data collection on this variable began after 2020); RBD Screening Questionnaire $n=3$ ; SCOPA-AUT $n=5$ .
<sup>a</sup> Comparisons across NSD stage at baseline used Chi-Square or Fisher's Exact tests for categorical variables and Kruskal-Wallis tests for continuous variables.
<sup>b</sup> No participants were categorized as Parkinson's Disease Dementia at baseline.

### Baseline clinical characteristics differ between NSD stages

Baseline disease characteristics of the NSD sporadic Parkinson’s Disease cohort are summarized in **Table 1**. Notably, participants recruited with the inclusion criteria of early untreated Parkinson’s Disease had a separation in NSD stages, with 67% (*n*=230) belonging to NSD Stage 3, 23% (*n*=79) NSD Stage 2b and 10% (*n*=35) NSD Stage 4. 275 (80%) of participants had an UPSIT score in the lower 15^th^ percentile expected for age and sex. As expected, there were stage dependent differences in MDS-UPDRS parts I, II as these were used as stage anchors, but also in other clinical measures across motor and non-motor domains, including MDS-UPDRS part III, as well as RBDSQ, GDS and SCOPA-AUT. While MoCA scores differed across NSD stage (*p*=0.041) with the lowest being the NSD Stage 3 group (27 [17-30]), there were no differences in cognitive categorization between groups.

### Baseline biologic characteristics of NSD participants

#### DAT binding measures

Participants in NSD Stage 4 had lower striatum binding compared to stages 2b and 3 (medians of 1.30 vs 1.52 and 1.33 respectively). Age/sex putaminal SBR was significantly different across groups, with lower scores in the stage 4 group compared to stages 2b and 3 (medians of 0.28 vs 0.35 and 0.30 respectively) **(Table 2).**

**Table 2.** Baseline Biological Characteristics.

|  | NSD Stage at Baseline |  |  |  |  |
| --- | --- | --- | --- | --- | --- |
|  | SAA +<br>(N = 344) | Stage 2b<br>(N = 79) | Stage 3<br>(N = 230) | Stage 4<br>(N = 35) | p-Value<sup>a</sup> |
| DAT binding Imaging Measures |  |  |  |  |  |
| Age/Sex-Expected Lowest Putamen SBR, Median (Min, Max) | 0.31 (0.06, 0.73) | 0.35 (0.18, 0.72) | 0.30 (0.06, 0.73) | 0.28 (0.10, 0.71) | 0.003 |
| Mean Striatum Binding, Median (Min, Max) | 1.36 (0.31, 2.59) | 1.52 (0.84, 2.59) | 1.33 (0.31, 2.52) | 1.30 (0.71, 2.36) | < 0.001 |
| Fluid Biomarkers |  |  |  |  |  |
| Serum NfL pg/mL, Median (Min, Max) | 11.4 (2.5, 76.6) | 11.5 (2.5, 46.6) | 11.3 (2.8, 32.5) | 13.5 (4.1, 76.6) | 0.639 |
| Urate mg/dL, Median (Min, Max) | 315.0 (167.0, 541.0) | 315.0 (172.0, 512.0) | 315.0 (167.0, 541.0) | 274.0 (202.0, 488.0) | 0.237 |
| CSF A-beta 1-42 pg/mL, Median (Min, Max) | 803.8 (249.0, 1467) | 831.6 (346.6, 1467) | 802.3 (249.0, 1419) | 669.4 (362.4, 1461) | 0.303 |
| CSF t-tau pg/mL, Median (Min, Max) | 157.3 (80.9, 467.0) | 154.3 (87.7, 290.1) | 157.3 (80.9, 467.0) | 175.0 (85.0, 302.9) | 0.615 |
| CSF p-tau pg/mL, Median (Min, Max) | 13.4 (8.0, 40.1) | 12.9 (8.0, 26.8) | 13.5 (8.0, 40.1) | 15.4 (8.2, 28.7) | 0.202 |
| CSF t-tau/A-beta 1-42 ratio, Median (Min, Max) | 0.18 (0.10, 0.72) | 0.18 (0.11, 0.39) | 0.18 (0.10, 0.69) | 0.19 (0.13, 0.72) | 0.276 |
| CSF p-tau/A-beta 1-42 ratio, Median (Min, Max) | 0.02 (0.01, 0.07) | 0.02 (0.01, 0.04) | 0.02 (0.01, 0.07) | 0.02 (0.01, 0.07) | 0.422 |
| CSF A-beta 1-42 ≤ 683 pg/mL (A+), n (%) | 108 (32%) | 23 (30%) | 69 (31%) | 16 (47%) | 0.141 |
| CSF p-tau ≥ 13 pg/mL (T+), n (%) | 166 (49%) | 35 (45%) | 112 (49%) | 19 (54%) | 0.678 |
| Serum NfL ≥ 19.05 pg/mL (N+), n (%) | 44 (14%) | 10 (14%) | 25 (12%) | 9 (30%) | 0.027 |
| Genetic Risk Score, Median (Min, Max) | -0.0094 (-0.0193, -0.0011) | -0.0097 (-0.0193, -0.0022) | -0.0093 (-0.0187, -0.0011) | -0.0092 (-0.0180, -0.0034) | 0.869 |

A-beta 1-42= Amyloid-beta 1-42, DAT= Dopamine transporter, NfL=Neurofilament light chain, NSD= Neuronal Synuclein Disease, p-tau= Tau phosphorylated at threonine 181 position, SBR=Striatal binding ratio, t-tau=total tau,
<sup>a</sup> Comparisons across NSD stage at baseline used Chi-Square or Fisher's Exact tests for categorical variables and Kruskal-Wallis tests for continuous variables.
Missing: Serum NfL *n*=32; Urate *n*=3; CSF A-beta 1-42 *n*=35; CSF t-tau *n*=17; CSF p-tau *n*=37; CSF t-tau/A-beta 1-42 ratio *n*=47; CSF p-tau/A-beta 1-42 ratio *n*=68; CSF A-beta 1-42 ≤ 683 *n*=7; CSF p-tau ≥ 13 *n*=4; Serum NfL ≥ 19.05 *n*=32; Genetic Risk Score *n*=6.

#### Fluid biomarkers

Baseline biologic characteristics of the NSD sporadic Parkinson’s Disease cohort presented by NSD stage are summarized in **Table 2**. There was no difference in median serum NfL across the groups but the percent of participants with serum NfL ≥ 19.05 pg/mL differed across baseline stage (*p*=0.027). The stage 4 group had the highest percentage of participants with elevated serum NfL (NSD Stage 2b 14%, Stage 3 12%, Stage 4 30%). Other fluid biomarkers did not show a difference between NSD stages **(Table 2).**

### Longitudinal characteristics

#### PPMI retention over time

Median follow up from baseline was 10.1 [0, 13.4] years. At 11 years, there were data available for 153 participants, which represents a 49% retention rate over study duration. 35 participants (10%) had died by the end of the follow-up period.

#### Clinical characteristics

As expected, motor clinical scores worsened over time. Mean [SD] MDS-UPDRS Part III medications ON score at year 11 was 28.4 [14.3], which represents a 36% increase compared to baseline OFF scores despite mean of 967.53 [501.93] LEDD use (96% on Parkinson’s Disease medication at year 11). Mean [SD] MDS-UPDRS Part IV score at year 11 was 4.5 [4]. As a group, mean modified S&E was 78.5% at 11 years. From a non-motor perspective, SCOPA-AUT scores also increased over time, with a mean [SD] 16 [7.5] and RBDSQ mean [SD] 5.6 [3.5] at year 11 **(Table 3)**. Reflecting the progression of functional impairment, there was a consistent increase in the percentage of individuals in NSD stage 4 and above during follow-up. By year 11 of *n=*153 participants, 44% were in NSD stage 3, another 44% had progressed to NSD stage 4, and 8% had advanced to NSD stage 5 **(Figure 1)**. Complete complement of longitudinal clinical data is presented in **Supplementary Table 3**.

**Figure 1.**
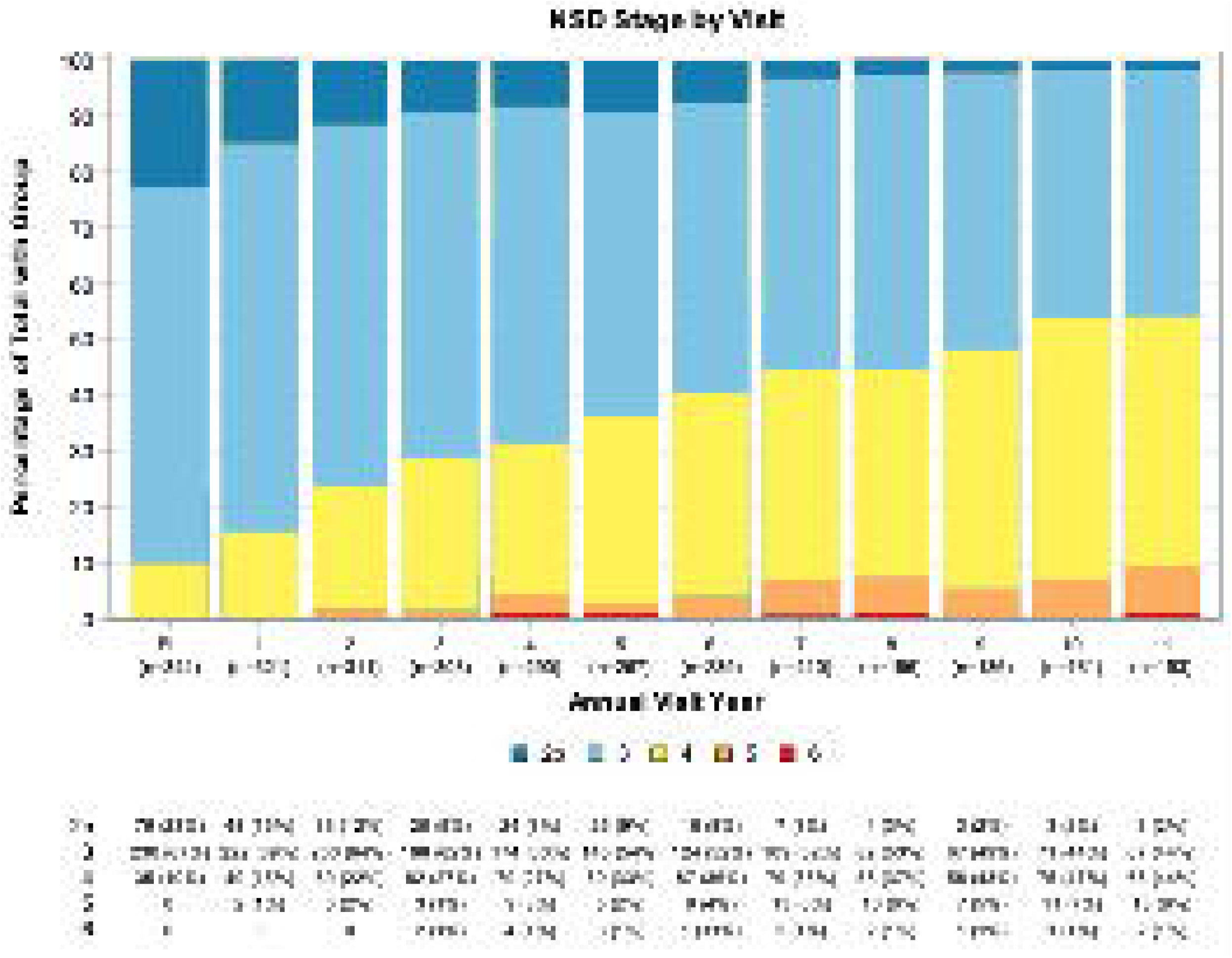
Neuronal Synuclein Stage (NSD) by visit. Shows the percentage of participants in each NSD Stage per annual visit over 11 years. Lower panel data shows total number of participants and percentage in each NSD Stage at each visit.

**Table 3.**
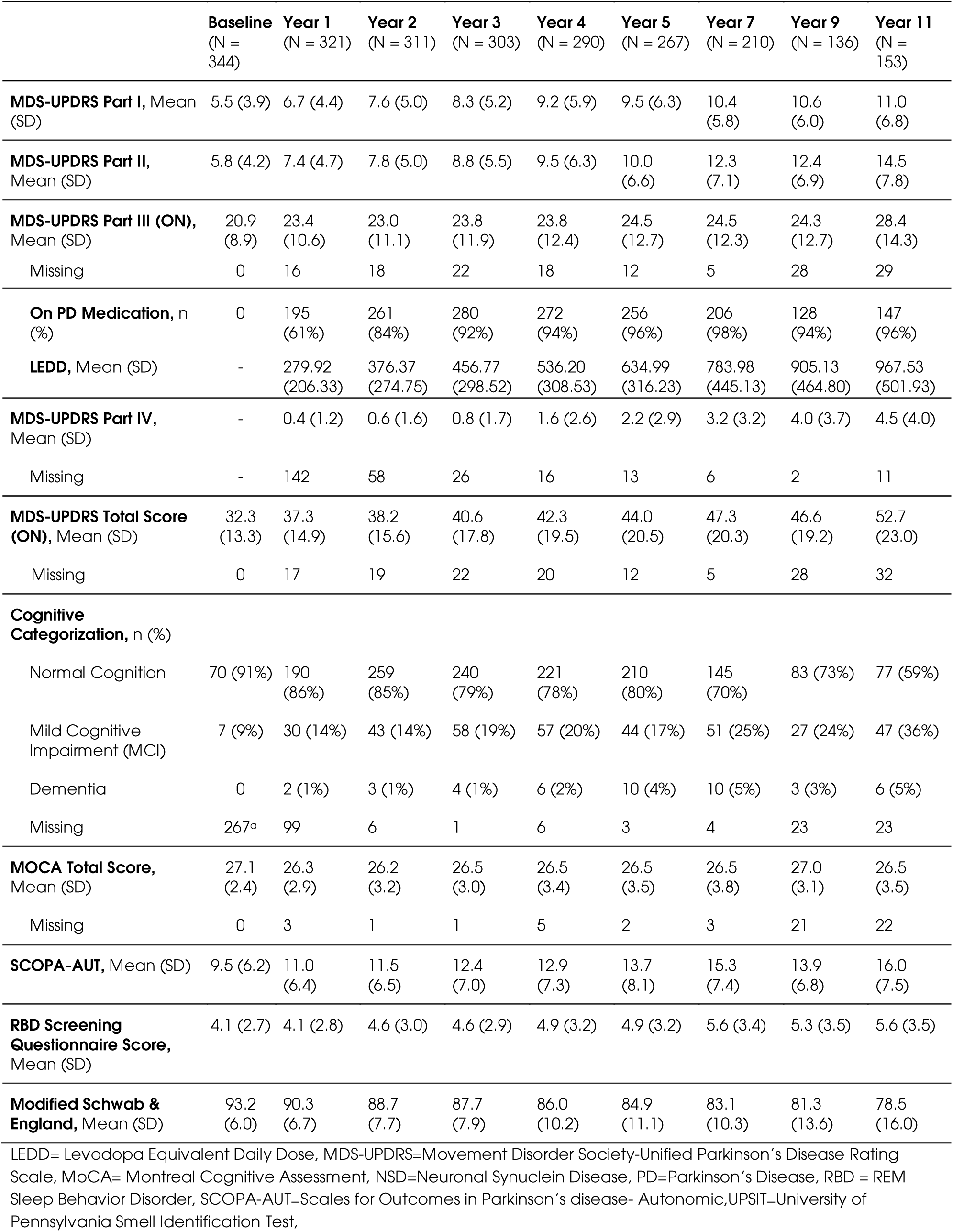
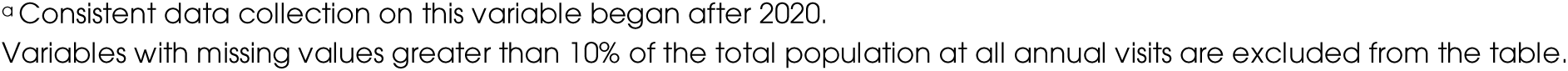
Longitudinal Clinical Characteristics.

|  | Baseline<br>(N = 344) | Year 1<br>(N = 321) | Year 2<br>(N = 311) | Year 3<br>(N = 303) | Year 4<br>(N = 290) | Year 5<br>(N = 267) | Year 7<br>(N = 210) | Year 9<br>(N = 136) | Year 11<br>(N = 153) |
| --- | --- | --- | --- | --- | --- | --- | --- | --- | --- |
| **MDS-UPDRS Part I, Mean (SD)** | 5.5 (3.9) | 6.7 (4.4) | 7.6 (5.0) | 8.3 (5.2) | 9.2 (5.9) | 9.5 (6.3) | 10.4 (5.8) | 10.6 (6.0) | 11.0 (6.8) |
| **MDS-UPDRS Part II, Mean (SD)** | 5.8 (4.2) | 7.4 (4.7) | 7.8 (5.0) | 8.8 (5.5) | 9.5 (6.3) | 10.0 (6.6) | 12.3 (7.1) | 12.4 (6.9) | 14.5 (7.8) |
| **MDS-UPDRS Part III (ON), Mean (SD)** | 20.9 (8.9) | 23.4 (10.6) | 23.0 (11.1) | 23.8 (11.9) | 23.8 (12.4) | 24.5 (12.7) | 24.5 (12.3) | 24.3 (12.7) | 28.4 (14.3) |
| Missing | 0 | 16 | 18 | 22 | 18 | 12 | 5 | 28 | 29 |
| **On PD Medication, n (%)** | 0 | 195 (61%) | 261 (84%) | 280 (92%) | 272 (94%) | 256 (96%) | 206 (98%) | 128 (94%) | 147 (96%) |
| **LEDD, Mean (SD)** | - | 279.92 (206.33) | 376.37 (274.75) | 456.77 (298.52) | 536.20 (308.53) | 634.99 (316.23) | 783.98 (445.13) | 905.13 (464.80) | 967.53 (501.93) |
| **MDS-UPDRS Part IV, Mean (SD)** | - | 0.4 (1.2) | 0.6 (1.6) | 0.8 (1.7) | 1.6 (2.6) | 2.2 (2.9) | 3.2 (3.2) | 4.0 (3.7) | 4.5 (4.0) |
| Missing | - | 142 | 58 | 26 | 16 | 13 | 6 | 2 | 11 |
| **MDS-UPDRS Total Score (ON), Mean (SD)** | 32.3 (13.3) | 37.3 (14.9) | 38.2 (15.6) | 40.6 (17.8) | 42.3 (19.5) | 44.0 (20.5) | 47.3 (20.3) | 46.6 (19.2) | 52.7 (23.0) |
| Missing | 0 | 17 | 19 | 22 | 20 | 12 | 5 | 28 | 32 |
| **Cognitive Categorization, n (%)** |  |  |  |  |  |  |  |  |  |
| Normal Cognition | 70 (91%) | 190 (86%) | 259 (85%) | 240 (79%) | 221 (78%) | 210 (80%) | 145 (70%) | 83 (73%) | 77 (59%) |
| Mild Cognitive Impairment (MCI) | 7 (9%) | 30 (14%) | 43 (14%) | 58 (19%) | 57 (20%) | 44 (17%) | 51 (25%) | 27 (24%) | 47 (36%) |
| Dementia | 0 | 2 (1%) | 3 (1%) | 4 (1%) | 6 (2%) | 10 (4%) | 10 (5%) | 3 (3%) | 6 (5%) |
| Missing | 267<sup>a</sup> | 99 | 6 | 1 | 6 | 3 | 4 | 23 | 23 |
| **MOCA Total Score, Mean (SD)** | 27.1 (2.4) | 26.3 (2.9) | 26.2 (3.2) | 26.5 (3.0) | 26.5 (3.4) | 26.5 (3.5) | 26.5 (3.8) | 27.0 (3.1) | 26.5 (3.5) |
| Missing | 0 | 3 | 1 | 1 | 5 | 2 | 3 | 21 | 22 |
| **SCOPA-AUT, Mean (SD)** | 9.5 (6.2) | 11.0 (6.4) | 11.5 (6.5) | 12.4 (7.0) | 12.9 (7.3) | 13.7 (8.1) | 15.3 (7.4) | 13.9 (6.8) | 16.0 (7.5) |
| **RBD Screening Questionnaire Score, Mean (SD)** | 4.1 (2.7) | 4.1 (2.8) | 4.6 (3.0) | 4.6 (2.9) | 4.9 (3.2) | 4.9 (3.2) | 5.6 (3.4) | 5.3 (3.5) | 5.6 (3.5) |
| **Modified Schwab & England, Mean (SD)** | 93.2 (6.0) | 90.3 (6.7) | 88.7 (7.7) | 87.7 (7.9) | 86.0 (10.2) | 84.9 (11.1) | 83.1 (10.3) | 81.3 (13.6) | 78.5 (16.0) |

LEDD= Levodopa Equivalent Daily Dose, MDS-UPDRS=Movement Disorder Society-Unified Parkinson's Disease Rating Scale, MoCA= Montreal Cognitive Assessment, NSD=Neuronal Synuclein Disease, PD=Parkinson's Disease, RBD = REM Sleep Behavior Disorder, SCOPA-AUT=Scales for Outcomes in Parkinson's disease- Autonomic, UPSIT=University of Pennsylvania Smell Identification Test,
<sup>a</sup> Consistent data collection on this variable began after 2020.
Variables with missing values greater than 10% of the total population at all annual visits are excluded from the table.

#### Biologic characteristics

DAT binding data were collected for four years. All DAT binding measures worsened over time, with mean (SD) age/sex expected lowest putamen SBR of 0.24 (0.09), mean (SD) striatum binding 1.03 (0.33), mean (SD) caudate binding 1.51 (0.51) and mean (SD) putamen binding 0.55 (0.19) at 4 years **(Table 4).**

**Table 4.** Longitudinal Imaging and Fluid Biomarker Characteristics.

|  | Baseline<br>(N = 344) | Year 1<br>(N = 321) | Year 2<br>(N = 311) | Year 3<br>(N = 303) | Year 4<br>(N = 290) | Year 5<br>(N = 267) |
| --- | --- | --- | --- | --- | --- | --- |
| **DAT binding Imaging Measures<sup>a</sup>** |  |  |  |  |  |  |
| **Age/Sex-Expected Lowest Putamen SBR, Mean (SD)** | 0.33 (0.11) | 0.29 (0.10) | 0.27 (0.10) | NA | 0.24 (0.09) | NA |
| **Mean Striatum Binding, Mean (SD)** | 1.41 (0.37) | 1.24 (0.34) | 1.16 (0.36) | NA | 1.03 (0.33) | NA |
| **Mean Caudate Binding, Mean (SD)** | 2.00 (0.53) | 1.80 (0.50) | 1.68 (0.53) | NA | 1.51 (0.51) | NA |
| **Mean Putamen Binding, Mean (SD)** | 0.81 (0.26) | 0.69 (0.22) | 0.64 (0.23) | NA | 0.55 (0.19) | NA |
| **Missing DAT** | 0 | 16 | 19 | NA | 42 | NA |
| **Fluid Biomarkers** |  |  |  |  |  |  |
| **Serum NfL pg/mL, Mean (SD)** | 13.0 (7.3) | 14.2 (10.9) | 15.0 (10.1) | 16.1 (9.9) | - | 18.5 (14.1) |
| Missing | 32 | 60 | 43 | 23 | - | 7 |
| **Urate mg/dL, Mean (SD)** | 317.0 (80.1) | 310.1 (74.9) | 309.2 (75.2) | 313.6 (76.6) | 309.8 (76.3) | 312.1 (79.9) |
| **CSF A-beta 1-42 pg/mL, Mean (SD)** | 826.5 (290.5) | 793.5 (284.1) | 808.0 (289.8) | 805.3 (278.0) | 784.5 (272.4) | 777.9 (286.1) |
| Missing | 35 | 77 | 79 | 95 | 117 | 119 |
| **CSF t-tau pg/mL, Mean (SD)** | 169.7 (58.3) | 165.6 (59.2) | 168.6 (64.1) | 169.5 (60.0) | 170.6 (67.7) | 162.0 (61.1) |
| Missing | 17 | 58 | 61 | 77 | 102 | 113 |
| **CSF p-tau pg/mL, Mean (SD)** | 14.9 (5.4) | 14.8 (5.2) | 14.9 (5.8) | 14.8 (5.4) | 14.5 (6.1) | 14.0 (5.4) |
| Missing | 37 | 87 | 85 | 93 | 112 | 119 |
| **CSF t-tau/A-beta 1-42 ratio, Mean (SD)** | 0.21 (0.09) | 0.21 (0.10) | 0.22 (0.11) | 0.21 (0.10) | 0.23 (0.14) | 0.22 (0.13) |
| Missing | 47 | 83 | 82 | 100 | 120 | 124 |
| **CSF p-tau/A-beta 1-42 ratio, Mean (SD)** | 0.02 (0.01) | 0.02 (0.01) | 0.02 (0.01) | 0.02 (0.01) | 0.02 (0.01) | 0.02 (0.01) |
| Missing | 68 | 112 | 106 | 116 | 130 | 130 |
| **CSF A-beta 1-42 ≤ 683 pg/mL (A+), n (%)** | 108 (32%) | 95 (35%) | 89 (35%) | 73 (32%) | 65 (34%) | 68 (43%) |
| Missing | 7 | 53 | 60 | 76 | 99 | 108 |
| **CSF p-tau ≥ 13 pg/mL (T+), n (%)** | 166 (49%) | 123 (46%) | 118 (47%) | 110 (48%) | 84 (44%) | 66 (42%) |
| Missing | 4 | 52 | 59 | 72 | 99 | 108 |
| **Serum NfL ≥ 19.05 pg/mL (N+), n (%)** | 44 (14%) | 44 (17%) | 59 (22%) | 72 (26%) | - | 82 (32%) |
| Missing | 32 | 60 | 43 | 23 | - | 7 |

A-beta 1-42= Amyloid-beta 1-42, DAT= Dopamine transporter, NfL=Neurofilament light chain, NSD= Neuronal Synuclein Disease, p-tau= Tau phosphorylated at threonine 181 position, SBR=Striatal binding ratio, t-tau=total tau, Variables with missing values less than 10% of the total population at all annual visits are excluded from the table.
<sup>a</sup> DAT obtained years 1, 2 and 4 only.

Data for different biofluid biomarkers are summarized in **Table 4**. CSF amyloid and tau markers varied over follow up, without a consistent trajectory over the 5 years of collected data. Serum NfL consistently increased over follow-up. The percentage of participants with a serum NfL ≥ 19.05 pg/mL also consistently increased over 5 years (32% by year 5).

#### A majority of NSD participants remain cognitively intact at 11 years

Of the 153 participants who had 11 years of follow-up, 59% (*n=*77) were categorized with normal cognition, 36% (*n*=47) were categorized as MCI and 5% (*n=*6) with Parkinson’s Disease. Likewise, the mean MoCA score was 26.5 (SD 3.5) at 11 years follow up **(Table 3).**

Time to investigator-determined development of mild cognitive impairment or dementia was significantly different when comparing baseline NSD stage 2b and 4. Median (95%CI) time was 11.93 (10.02, 12.01) years for participants presenting as NSD Stage 2b, 7.19 (6.13, 8.98) years for NSD Stage 3 and 3.10 (1.97, 4.99) years for participants in NSD Stage 4. The probability of reaching investigator-determined cognitive decline was higher in NSD stage 4 participants compared to those in NSD stage 2b (hazards ratio [HR]: 3.64, 95% CI 2.07, 6.40, *p*=<0.001; (**Figure 2D**). Even after accounting for the competing risks of death and informative withdrawal in the sensitivity analysis, those in NSD Stage 4 had a significantly higher probability of reaching this outcome compared to those in NSD Stage 2b, with a HR of 2.73 (95% CI 1.45, 5.15, *p=*0.0019) **(Supplementary Table 4).**

**Figure 2.**
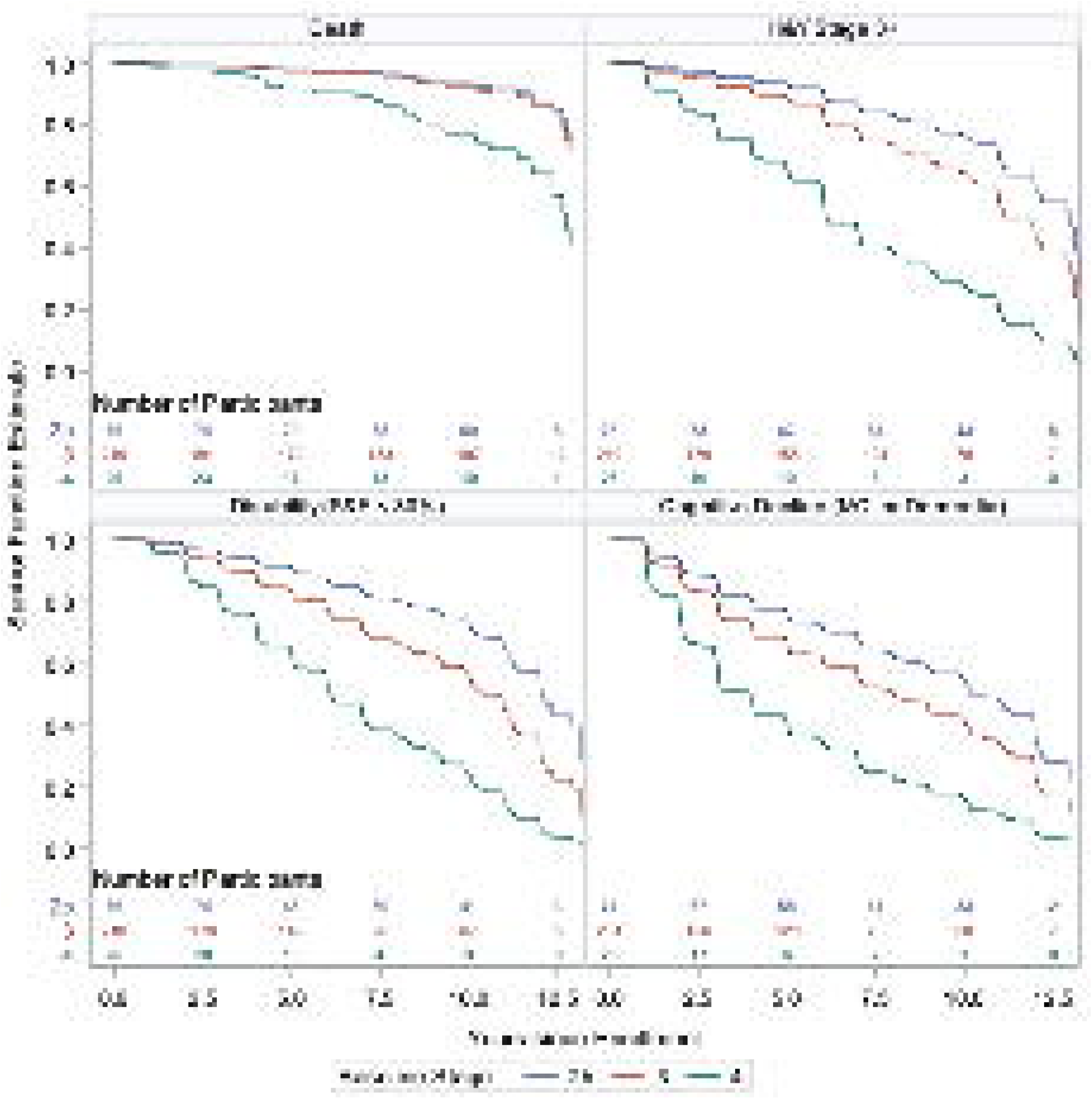
Time to death, postural instability, disability and cognitive decline by NSD Stage, adjusted by age, sex, and education. Survival estimates are presented for each NSD Stage. NSD Stage 4 reached all milestones earlier compared to Stage 2b (blue line). **(A)** Death (HR 4.58 [1.32,15.93], *p=*0.016) **(B)** Postural instability (Hoehn & Yahr ≥3) (HR 6.69 [3.3,13.58], *p* <0.0001)**, (C)** Disability (Schwab & England < 80%) (HR 4.87 [2.55,9.28], *p* <0.0001), and **(D)** Cognitive decline (investigator determined Mild cognitive impairment or Parkinson’s Disease dementia) (HR 3.64 [2.07,6.40], *p* <0.0001).

### Baseline NSD Stage as a predictor of progression

#### NSD Stage predicts time to disability

Median (95%CI) time to disability, defined by S&E score <80% was 11.98 (11.08, Inf) years for participants presenting as NSD Stage 2b; 10.29 (9.24, 11.10) years for NSD Stage 3 and 6.15 (3.89, 7.84) years for NSD Stage 4 participants. NSD Stage 3 and 4 participants had a higher probability of reaching a S&E score <80% compared to NSD Stage 2b (HR 1.88, 95%CI 1.21, 2.92, *p=*0.005; HR 4.87, 95%CI 2.55, 9.28, *p*= <0.001) **(Figure 2B).** Participants in NSD Stage 4 also had higher probability of reaching a H&Y ≥ 3 compared to NSD Stage 2b (HR 6.69, 95%CI 3.30, 13.58, *p=<*0.001) **(Figure 2C).** After accounting for death and informative withdrawal in the sensitivity analysis, the hazards of reaching both outcomes were significantly higher for those in NSD stage 4 compared to those in NSD stage 2b (HR 2.97, 95%CI 1.50, 5.88, *p* = 0.0018; HR 3.36, 95%CI 1.60, 7.06, *p =* 0.0014.(**Supplementary Table 4**).

#### NSD Stage predicts time to clinically meaningful milestones

Baseline NSD Stage was associated with time to reach every disease milestone, with participants in NSD Stage 4 reaching most disease milestones earlier **(Figure 3 A-E).** The sensitivity analysis accounting for death and informative withdrawal yielded the same conclusions, with the exception of the ‘motor complications’ milestone **(Supplementary Table 4)**.

**Figure 3.**
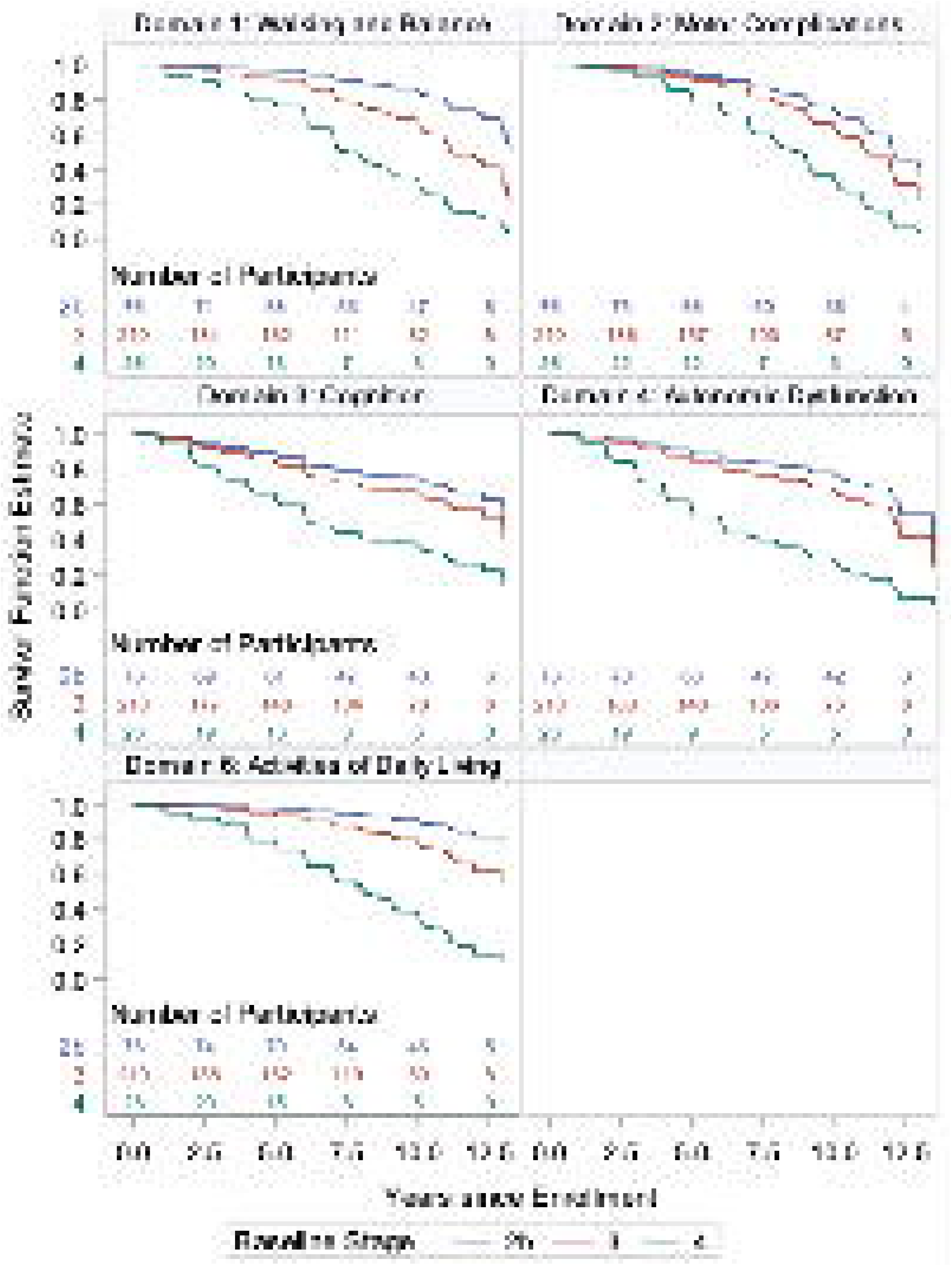
Time to milestone domains, by NSD Stage, adjusted by age, sex, and education. Survival estimates are presented for each NSD Stage. NSD Stage 4 (green line) reached all milestones earlier compared to NSD Stage 2b (blue line). **(A)** Domain 1: Walking and Balance (HR 8.26 [3.82, 17.9], *p*<0.0001)**, (B)** Domain 2: Motor Complications (HR 3.63 [1.70, 7.74], *p*=0.0009)**, (C)** Domain 3: Cognition (HR 3.96 [2.01, 7.79], *p*<0.0001), **(D)** Domain 4: Autonomic Dysfunction (HR 5.79 [2.78, 12.08], *p*<0.0001), **(E)** Domain 6: Activities of Daily Living (HR 11.03 [4.48, 27.14], *p*<0.0001). Domain 5: Functional dependence is not shown since it is defined by S&E <80% which is displayed in Figure 2.

#### Baseline NSD Stage is not associated with survival

Although survival time was reduced in NSD Stage 4 participants compared to those in NSD Stage 2b (**Figure 2A**), this difference was not statistically significant There was no significant difference in time to death between participants in NSD Stages 3 and 2b.

## Discussion

These data represent the first report of long-term outcomes (> 10 years) of participants with sporadic Parkinson’s Disease fulfilling criteria for NSD. There are several observations and conclusions that inform the field. Most importantly, our data demonstrate a much more optimistic outlook on Parkinson’s Disease progression compared to previously published and referenced data.^8–10^ The known mortality rate was 10%, and the majority of the retained participants maintained good functional status and remained cognitively intact. While these conclusions must be interpreted considering selective nature of participants generally recruited into studies, who might not be representative of the Parkinson’s Disease population at large, the same limitations apply to prior studies, such as the Sydney cohort^8–10^. These better outcomes might be a reflection of better comprehensive treatment paradigms available to Parkinson’s Disease patients today and could also be a reflection of a more biologically homogeneous population in our study. Compared to the Sydney cohort 10 year outcomes,^9^ our results are consistent with the data on long term cognitive outcomes in the PPMI cohort published by Gallagher *et al*^11^ and expand beyond the cognitive domain.

### NSD Stage differs at baseline in participants clinically diagnosed as sporadic Parkinson’s Disease

This presents the first analysis of sporadic Parkinson’s Disease participants who were defined and staged according to the NSD-ISS. The NSD is based on a biologic characterization of individuals, i.e. presence of abnormal α-syn (currently defined as a positive CSF-SAA), as well as dopamine deficiency (currently defined as DAT binding deficiency).^18^ NSD-ISS functional characterization is anchored on clinical scales commonly used in clinical practice. At baseline, despite identical inclusion criteria used for PPMI enrollment, participants represented a range of NSD stages. This is of significance since all participants were considered to be “early” Parkinson’s Disease, that is, within two years of clinical diagnosis, and treatment naïve. Even more, motor, non-motor and DAT binding characteristics significantly differed between participants in different stages at baseline. Our findings suggest that the arbitrary use of time since diagnosis does not clearly represent the underlying disease severity of individuals. This is of importance given that time since diagnosis is a common inclusion criterion for most clinical trials. As such, our data support the notion that the NSD-ISS could be used as a framework for selecting potential clinical trial candidates.

### NSD Stage as a predictor of disease progression

The results of this analysis support the validity of NSD-ISS as a predictor for disease progression. Prediction of disease and disease course is a critical challenge that influences patient counseling, care, treatment, and research. Within Parkinson’s Disease, meeting this challenge would allow appropriate planning for patients and symptom-specific care. These prediction tools would also facilitate more efficient execution of clinical trials.

Past attempts at the characterization of disease subtypes have relied on clinical measures, separating the disease into early-onset versus late-onset (based on age at time of diagnosis), slowly-progressing “benign” versus fast-progressing “malignant” subtypes, Parkinson’s Disease with or without dementia, or into a tremor-dominant versus a postural instability with gait disorder subtypes.^4,43^ However, these models have shown instability over time.^44^ A past study employed cluster analysis to classify patient subtypes and ascertain their respective progression rates, but it was limited to data from just two time points.^45^ Some models have identified individual variables correlating with progression, often being restricted to a narrow set of predictors.^45^, ^Macleod, 2018^ ^#49^ For example, the Parkinson’s Disease and parkinsonism in North East Scotland (PINE) and ParkWest studies highlighted older age, male sex, and greater severity of axial features as predictors of mortality, with additional factors such as smoking history and cognitive assessments linked to loss of independence.^46^ Modeling paradigms involving the development of composite scores or the use of machine learning algorithms to identify disease subtypes have also been proposed.^45,47–51^ Nonetheless, the depth of phenotypic information and longitudinal assessments in these studies were variable and often limited to certain clinical features and short-term follow-up. These models also present challenges for implementation in clinical settings. Recently, the Movement Disorder Society Task Force on Subtyping published their critical evaluation on current subtyping systems and concluded that the subtyping studies undertaken to date have significant methodologic shortcomings, with questionable clinical applicability and with unknown biological relevance.^52^ In contrast, the NSD-ISS offers a straightforward and practical model that correlates with both motor and non-motor disease progression, providing a comprehensive and consistent association across the full disease spectrum. Currently, the NSD-ISS is a research framework that can enhance design and implementation of clinical trials by reducing baseline heterogeneity of the participants. In the future it could serve an important role of assisting baseline prognostication in the clinical setting.

### Comparison to other longitudinal Parkinson’s Disease cohorts

We compared our data to some of the most relevant longitudinal cohorts described to date. The largest past longitudinal cohort studies in Parkinson’s Disease include the Sydney Multicenter Study of Parkinson’s Disease,^8–10^ the Cambridgeshire Parkinson’s Incidence from GP to Neurologist (CamPaIGN),^53–55^ and the PINE^56,57^ studies. The Sydney Multicenter Study followed *n=*149 Parkinson’s Disease participants randomized to low-level levodopa versus low-dose bromocriptine over 20 years. By 20 years follow up, *n=*30 participants were alive and of those, *n=*25 had dementia^10^. The CamPaIGN and PINE studies represent an unselected population-representative incident cohort clinically diagnosed with Parkinson’s Disease (*n*=142 and *n*=199 respectively) followed for 13 years and 9.5 years respectively. These cohorts describe most participants presenting with disability and dementia by 10 years follow up. Nonetheless, none of these cohorts had biological characterization or identify models to predict subgroups who may reach these endpoints earlier. In this way, the PPMI cohort represents the largest phenotypically and biologically characterized cohort of participants clinically diagnosed as Parkinson’s Disease, who have been longitudinally followed for over 10 years.

Postural instability was the first milestone reached by NSD participants, which is in line with what was described in the CamPaign and Sydney cohorts. Nonetheless, median time to Hoehn & Yahr ≥ 3 was longer in PPMI (6.0 years among NSD Stage 4) compared to the CamPaign (4.7 years)^58^ or Sydney cohorts (3.5 years).^9^ Time to disability was also longer in the PPMI cohort (6.2 years among NSD Stage 4) compared to the PINE study, where 50% of those with Parkinson’s Disease were dead or dependent three years from diagnosis.^59^ Finally, mortality in the PPMI study was lower compared to prior studies, with a cumulative probability of survival of around 75% at 10 years for participants in NSD Stage 4. In the Sydney study, 38% of participants had died at 10 years.^8^ In the CamPaIGN study, at 10 years, survival analysis indicated a cumulative probability of survival of 45%.^58^ While these differences could be due to a younger age in the PPMI cohort and participant selection, it could also reflect analyzing a biologically homogeneous population, since participants with parkinsonism associated with non-synuclein pathology have a more aggressive course of the disease.^60,61^ Lastly, it could also represent a change in the natural history of the disease in contemporary cohorts, with a more optimistic outlook for patients.

In line with this, the prevalence of dementia is significantly lower compared to what has been described in previous cohorts. In the Sydney study, at 20 years, 83% of survivors (*n=*36) fulfilled criteria for dementia.^10^ The CamPaiGN cohort showed an increase in dementia incidence with disease duration and described a probability of 46% at 10-years, with higher baseline UPDRS motor score being predictive of dementia.^58^ This is in line with what has been described for other longitudinal studies following Parkinson’s Disease cohorts from time of diagnosis.^62,63^ In the PPMI cohort, only 36% of NSD participants presented MCI at 10 years and only 4% met criteria for dementia. Interestingly, despite all participants presenting with normal cognition at baseline, when looking at individual NSD stages, participants belonging to NSD Stage 4 reached a categorization of MCI or dementia in a median time of 3.1 years. Among those with NSD stage 4 at baseline, the estimated 10-year probability of reaching MCI or dementia was 80%, which is in line with what was described in the Sydney multicenter cohort.^10^ This suggests that, while dementia may not be inevitable for all NSD participants, those with NSD Stage 4 at baseline have a higher risk of developing cognitive changes, again supporting NSD-ISS as a tool for baseline prediction of progression.

### DAT binding as an imaging biomarker

Our analysis showed that striatal binding ratio and age and sex adjusted values were significantly lower at baseline and remained lower over follow up in participants in NSD-Stage 4. The degree of DAT binding has been described to be associated with worsened longitudinal motor scores in prior, smaller studies, though the data are inconsistent. An analysis of a subset of the Anti α-Synuclein Antibody in Early Parkinson’s Disease (PASADENA) study data showed that ipsilateral putamen DAT binding can predict progression of motor signs in early Parkinson’s Disease, albeit this was a small data set (*n*=76 participants) with data available for 12 months.^64^ The Parkinson Associated Risk Syndrome study showed that prodromal participants with hyposmia and with dopamine transporter deficit at baseline were at a higher risk of developing the motor symptoms of Parkinson’s Disease.^65^ Nonetheless, other studies have failed to show these associations with progression. Our data lays the groundwork for utilizing DAT binding as a potential marker for disease progression. However, one limitation is availability of PPMI DAT binding data is restricted to the first four years of follow-up. To fully assess the utility of DAT binding measures over the course of NSD, extended longitudinal data are required.

### Longitudinal change in fluid biomarkers of neurodegeneration

We did not identify a reliable fluid biomarker that demonstrated longitudinal quantitative change at the group level. An obvious limitation and area for future research is the development of a quantitative biomarker of α-syn pathology and assessing if it will correlate with longitudinal changes in clinical measures. There has been significant interest in biomarkers such as Aβ 1–42, tau, and p-tau for providing insights into the progression of Parkinson’s Disease, particularly in evaluating cognitive decline and motor function. However, no consistent results have been reported thus far.^66–68^ In our study, we also observed no consistent trajectory of CSF-Alzheimer’s Disease biomarkers over time, and these biomarkers did not differ between NSD stages at baseline. There has been a lot of interest in NfL as a biomarker of neurodegeneration across neurological diseases. Plasma NfL concentration reflects neuro-axonal damage and has been described as a marker for ongoing neurodegeneration.^69–71^ In this analysis, a higher proportion of participants in NSD Stage 4 at baseline had a serum NfL level ≥ 19.05 pg/mL. Additionally, data available over 5 years shows a consistent increase in serum NfL levels in NSD participants as a group. Mollenhauer *et al*^71^have reported higher mean baseline serum NfL in Parkinson’s Disease compared to healthy controls, with levels increasing over time; this group also reported on an association with motor scores. Another study, which included “late-stage” Parkinson’s Disease participants (defined as disease duration greater than 5 years), described higher serum NfL in those presenting with hallucinations, dementia or recurrent falls.^72^ However, a limitation of this analysis is that data were only available for the first 5 years of follow up. Ongoing efforts at PPMI to collect long-term broader spectrum of biological data will be crucial for further understanding of the disease biology.

### Limitations and future work

The main methodological concern in longitudinal studies of this type is attrition. Participant retention in PPMI is lower compared to prior longitudinal cohorts.^8–10,56–58^ which may reflect the duration and scope of ascertainments in PPMI. In this analysis, we are particularly concerned with understanding any bias introduced by participants who drop out early due to worsening Parkinson’s Disease symptoms. We include the sensitivity analysis considering both death and informative dropout as competing events in an attempt to assess the impact of attrition due to worsening symptoms of Parkinson’s Disease on our analysis results. Informative dropout was categorized conservatively to include anyone who did not have a reported operational (not related to Parkinson’s Disease progression) reason for withdrawal from the study. The results of this analysis yield attenuated results but still support the conclusion that NSD stage, comparing 2b and 4 at baseline, are predictors of progression outcomes.

The event-based model, as currently designed, presumes a uniform progression sequence across NSD participants, which is not reflective of the condition’s well-documented variability in clinical presentation. Future research will focus on refining this model to better differentiate the timing and sequence of progression events.

Generalizability might be limited by the characteristics of the PPMI cohort. Notably, the PPMI group is considerably younger and with a lower LEDD at baseline compared to incident enrolled cohorts, such as CamPAIGN or PINE. Nonetheless, the PPMI cohort is comparable to other clinical trial cohorts,^73,74^ as such these results are most relevant to the design and planning of future clinical trials. We also acknowledge that the PPMI cohort lacks racial and ethnic diversity. As such, there are efforts underway to increase the enrollment of traditionally underrepresented groups into this study.

Additionally, NSD criteria necessitates the presence of synucleinopathy, currently assessed as a positive CSF-SAA marker. We recognize the need for less invasive methods to assess SAA in more readily accessible biofluids to facilitate broader application. Despite this, our findings affirm the utility of the NSD-ISS within a research context, particularly for clinical trials.

While PPMI is focused on biological characterization of the participants, current data fails to identify a quantitative biomarker of NSD progression. PPMI is committing substantial effort to develop a quantitative biomarker of α-syn pathology and to incorporate and assess multiple novel biomarkers as such become available. We have plans for multiple lines of future work to further improve understanding of NSD participants. We concentrated on previously described biologic biofluids of interest. Nonetheless, other biologic measures, including omics data could be explored in the future, to further characterize biologic differences between stages. Additionally, we have focused on NSD participants enrolled in the Parkinson’s Disease cohort. Understanding the biology and progression of the SAA negative subset of participants in the sporadic Parkinson’s Disease cohort is of high priority and these data will be reported separately.

In conclusion, we present 11-year longitudinal follow up data on sporadic Parkinson’s Disease participants who fulfill criteria for NSD. We demonstrate that, at time of enrollment, there is significant heterogeneity of the participants recruited into the study under the currently used criteria of early Parkinson’s Disease. Baseline NSD stage predicted progression to clinically meaningful milestones, emphasizing the need for new clinical trial designs to shift from traditional, clinically based enrollment criteria and arbitrary time since diagnosis to a more precise biological and functional characterization of individuals. Additionally, we observed more favorable long-term outcomes—including survival, disability, postural instability, and cognitive decline—reflective of a contemporary observational study cohort. These findings provide a more hopeful perspective on the long-term progression of Parkinson’s Disease.

### Data availability

Data used in the preparation of this article were obtained on June 24th, 2024 from the PPMI database (www.ppmi-info.org/access-data-specimens/download-data), RRID:SCR 006431. For up-to-date information on the study, visit www.ppmi-info.org. This analysis was conducted by the PPMI Statistics Core and used actual dates of activity for participants, a restricted data element not available to public users of PPMI data. Access can be obtained upon request. Statistical analysis codes used to perform the analyses in this article are shared on Zenodo (10.5281/zenodo.11660808).

## Acknowledgments

PPMI, a public-private partnership, is funded by the Michael J. Fox Foundation for Parkinson’s Research and funding partners, including 4D Pharma, Abbvie, AcureX, Allergan, Amathus Therapeutics, Aligning Science Across Parkinson’s, AskBio, Avid Radiopharmaceuticals, BIAL, Biogen, Biohaven, BioLegend, BlueRock Therapeutics, Bristol-Myers Squibb, Calico Labs, Celgene, Cerevel Therapeutics, Coave Therapeutics, DaCapo Brainscience, Denali, Edmond J. Safra Foundation, Eil Lilly, GE HealthCare, Genentech, GSK, Golub Capital, Gain Therapeutics, Handl Therapeutics, Insitro, Janssen Neuroscience, Lundbeck, Merck, Meso Scale Discovery, Mission Therapeutics, Neurocrine Biosciences, Pfizer, Piramal, Prevail Therapeutics, Roche, Sanofi, Servier, Sun Pharma Advanced Research Company, Takeda, Teva, UCB, Vanqua Bio, Verily, Voyager Therapeutics, the Weston Family Foundation and Yumanity Therapeutics.

## Supplementary material

Supplementary material is available at *Brain* online

## Appendix 1

### PPMI study teams/Cores/collaborators for publications

#### Executive Steering Committee

Kenneth Marek, MD^1^ (Principal Investigator); Caroline Tanner, MD, PhD^8^; Tanya Simuni, MD^3^; Andrew Siderowf, MD, MSCE^11^; Douglas Galasko, MD^26^; Lana Chahine, MD^38^; Christopher Coffey, PhD^4^; Kalpana Merchant, PhD^58^; Kathleen Poston, MD^37^; Roseanne Dobkin, PhD^40^; Tatiana Foroud, PhD^14^; Brit Mollenhauer, MD^7^; Dan Weintraub, MD^11^; Ethan Brown, MD^8^; Karl Kieburtz, MD, MPH^22^; Mark Frasier, PhD^5^; Todd Sherer, PhD^5^; Sohini Chowdhury, MA^5^; Roy Alcalay, MD^45^ and Aleksandar Videnovic, MD^44^

#### Steering Committee

Duygu Tosun-Turgut, PhD^8^; Werner Poewe, MD^6^; Susan Bressman, MD^13^; Jan Hammer^14^; Raymond James, RN^21^; Ekemini Riley, PhD^39^; John Seibyl, MD^1^; Leslie Shaw, PhD^11^; David Standaert, MD, PhD^17^; Sneha Mantri, MD, MS^59^; Nabila Dahodwala, MD^11^; Michael Schwarzschild^44^; Connie Marras^42^; Hubert Fernandez, MD^24^; Ira Shoulson, MD^22^; Helen Rowbotham^2^; Paola Casalin^10^ and Claudia Trenkwalder, MD^7^

#### Michael J. Fox Foundation (Sponsor)

Todd Sherer, PhD; Sohini Chowdhury, MA; Mark Frasier, PhD; Jamie Eberling, PhD; Katie Kopil, PhD; Alyssa O’Grady; Maggie McGuire Kuhl; Leslie Kirsch, EdD and Tawny Willson, MBS

#### Study Cores, Committees and Related Studies: *(Include as applicable to the paper)*

*Project Management Core:* Emily Flagg, BA^1^

*Site Management Core:* Tanya Simuni, MD^3^; Bridget McMahon, BS^1^

Strategy and Technical Operations: Craig Stanley, PhD^1^; Kim Fabrizio, BA^1^

*Data Management Core:* Dixie Ecklund, MBA, MSN^4^; Trevis Huff, BSE^4^

*Screening Core:* Tatiana Foroud, PhD^14^; Laura Heathers, BA^14^; Christopher Hobbick, BSCE^14^; Gena Antonopoulos, BSN^14^

*Imaging Core:* John Seibyl, MD^1^; Kathleen Poston, MD^37^

*Statistics Core*: Christopher Coffey, PhD^4^; Chelsea Caspell-Garcia, MS^4^; Michael Brumm, MS^4^

*Bioinformatics Core*: Arthur Toga, PhD^9^; Karen Crawford, MLIS^9^

*Biorepository Core:* Tatiana Foroud, PhD^14^; Jan Hamer, BS^14^

*Biologics Review Committee*: Brit Mollenhauer^7^; Doug Galasko^26^; Kalpana Merchant^58^

*Genetics Core:* Andrew Singleton, PhD^12^

*Pathology Core:* Tatiana Foroud, PhD^14^; Thomas Montine, MD, PhD^37^

*Found:* Caroline Tanner, MD PhD^8^

*PPMI Online:* Carlie Tanner, MD PhD^8^; Ethan Brown, MD^8^; Lana Chahine, MD^38^; Roseann Dobkin, PhD^40^; Monica Korell, MPH^8^

#### Site Investigators

Charles Adler, PhD^48^; Roy Alcalay, MD^34^; Amy Amara, PhD^49^; Paolo Barone, PhD^29^; Bastiaan Bloem, PhD^57^ Susan Bressman, MD^15^; Kathrin Brockmann, MD^25^; Norbert Brüggemann, MD^56^; Lana Chahine, MD^38^; Kelvin Chou, MD^41^; Nabila Dahodwala, MD^11^; Alberto Espay, MD^31^; Stewart Factor, DO^15^; Hubert Fernandez, MD^24^; Michelle Fullard, MD^49^; Douglas Galasko, MD^26^; Robert Hauser, MD^18^; Penelope Hogarth, MD^16^; Shu-Ching Hu, PhD^20^; Michele Hu, PhD^55^; Stuart Isaacson, MD^30^; Christine Klein, MD^56^; Rejko Krueger, MD^2^; Mark Lew, MD^46^; Zoltan Mari, MD^53^; Connie Marras, PhD^42^; Maria Jose Martí, PhD^32^; Nikolaus McFarland, PhD^51^; Tiago Mestre, PhD^43^; Brit Mollenhauer, MD^7^; Emile Moukheiber, MD^27^; Alastair Noyce, PhD^60^; Wolfgang Oertel, PhD^61^; Njideka Okubadejo, MD^62^; Sarah O’Shea, MD^36^; Rajesh Pahwa, MD^45^; Nicola Pavese, PhD^54^; Werner Poewe, MD^6^; Ron Postuma, MD^52^; Giulietta Riboldi, MD^50^; Lauren Ruffrage, MS^17^; Javier Ruiz Martinez, PhD^33^; David Russell, PhD^1^; Marie H Saint-Hilaire, MD^21^; Neil Santos, BS^48^; Wesley Schlett^44^; Ruth Schneider, MD^22^; Holly Shill, MD^47^; David Shprecher, DO^23^; Tanya Simuni, MD^3^; David Standaert, PhD^17^; Leonidas Stefanis, PhD^35^; Yen Tai, PhD^28^; Caroline Tanner, PhD^8^; Arjun Tarakad, MD^19^; Eduardo Tolosa PhD^32^ and Aleksandar Videnovic, MD^44^

#### Coordinators

Susan Ainscough, BA^29^; Courtney Blair, MA^17^; Erica Botting^18^; Isabella Chung, BS^53^; Kelly Clark^23^; Ioana Croitoru^33^; Kelly DeLano, MS^31^; Iris Egner, PhD^6^; Fahrial Esha, BS^50^; May Eshel, MSc^34^; Frank Ferrari, BS^41^; Victoria Kate Foster^54^; Alicia Garrido, MD^32^; Madita Grümmer^56^; Bethzaida Herrera^47^; Ella Hilt^25^; Chloe Huntzinger, BA^49^; Raymond James, BS^21^; Farah Kausar, PhD^8^; Christos Koros, MD, PhD^35^; Yara Krasowski, MSc^57^; Dustin Le, BS^16^; Ying Liu, MD^49^; Taina M. Marques, PhD^2^; Helen Mejia Santana, MA^36^; Sherri Mosovsky, MPH^38^; Jennifer Mule, BS^24^; Philip Ng, BS^42^; Lauren O’Brien^45^; Abiola Ogunleye, PGDip^28^; Oluwadamilola Ojo, MD^62^; Obi Onyinanya, BS^27^; Lisbeth Pennente, BA^30^; Romina Perrotti^52^; Michael Pileggi, MS^52^; Ashwini Ramachandran, MSc^11^; Deborah Raymond, MS^13^; Jamil Razzaque, MS^55^; Shawna Reddie, BA^43^; Kori Ribb, BSN,^27^; Kyle Rizer, BA^51^; Janelle Rodriguez, BS^26^; Stephanie Roman, HS^1^; Clarissa Sanchez, MPH^19^; Cristina Simonet, PhD^28^; Anisha Singh, BS^22^; Elisabeth Sittig, RN^61^; Barbara Sommerfeld MSN^15^; Angela Stovall, BS^41^; Bobbie Stubbeman, BS^31^; Alejandra Valenzuela, BS^46^; Catherine Wandell, BS^20^; Diana Willeke^7^; Karen Williams, BA^3^ and Dilinuer Wubuli, MB^42^

Partners Scientific Advisory Board (Acknowledgement)

Funding: PPMI – a public-private partnership – is funded by the Michael J. Fox Foundation for Parkinson’s Research and funding partners, including 4D Pharma, Abbvie, AcureX, Allergan, Amathus Therapeutics, Aligning Science Across Parkinson’s, AskBio, Avid Radiopharmaceuticals, BIAL, BioArctic, Biogen, Biohaven, BioLegend, BlueRock Therapeutics, Bristol-Myers Squibb, Calico Labs, Capsida Biotherapeutics, Celgene, Cerevel Therapeutics, Coave Therapeutics, DaCapo Brainscience, Denali, Edmond J. Safra Foundation, Eli Lilly, Gain Therapeutics, GE HealthCare, Genentech, GSK, Golub Capital, Handl Therapeutics, Insitro, Jazz Pharmaceuticals, Johnson & Johnson Innovative Medicine, Lundbeck, Merck, Meso Scale Discovery, Mission Therapeutics, Neurocrine Biosciences, Neuron23, Neuropore, Pfizer, Piramal, Prevail Therapeutics, Roche, Sanofi, Servier, Sun Pharma Advanced Research Company, Takeda, Teva, UCB, Vanqua Bio, Verily, Voyager Therapeutics, the Weston Family Foundation and Yumanity Therapeutics.

1. Institute for Neurodegenerative Disorders, New Haven, CT

2. University of Luxembourg, Luxembourg

3. Northwestern University, Chicago, IL

4. University of Iowa, Iowa City, IA

5. The Michael J. Fox Foundation for Parkinson’s Research, New York, NY

6. Innsbruck Medical University, Innsbruck, Austria

7. Paracelsus-Elena Klinik, Kassel, Germany

8. University of California, San Francisco, CA

9. Laboratory of Neuroimaging (LONI), University of Southern California

10. BioRep, Milan, Italy

11. University of Pennsylvania, Philadelphia, PA

12. National Institute on Aging, NIH, Bethesda, MD

13. Mount Sinai Beth Israel, New York, NY

14. Indiana University, Indianapolis, IN

15. Emory University of Medicine, Atlanta, GA

16. Oregon Health and Science University, Portland, OR

17. University of Alabama at Birmingham, Birmingham, AL

18. University of South Florida, Tampa, FL

19. Baylor College of Medicine, Houston, TX

20. University of Washington, Seattle, WA

21. Boston University, Boston, MA

22. University of Rochester, Rochester, NY

23. Banner Research Institute, Sun City, AZ

24. Cleveland Clinic, Cleveland, OH

25. University of Tübingen, Tübingen, Germany

26. University of California, San Diego, CA

27. Johns Hopkins University, Baltimore, MD

28. Imperial College of London, London, UK

29. University of Salerno, Salerno, Italy

30. Parkinson’s Disease and Movement Disorders Center, Boca Raton, FL

31. University of Cincinnati, Cincinnati, OH

32. Hospital Clinic of Barcelona, Barcelona, Spain

33. Hospital Universitario Donostia, San Sebastian, Spain

34. Tel Aviv Sourasky Medical Center, Tel Aviv, Israel

35. National and Kapodistrian University of Athens, Athens, Greece

36. Columbia University Irving Medical Center, New York, NY

37. Stanford University, Stanford, CA

38. University of Pittsburgh, Pittsburgh, PA

39. Center for Strategy Philanthropy at Milken Institute, Washington D.C.

40. Rutgers University, Robert Wood Johnson Medical School, New Brunswick, New Jersey

41. University of Michigan, Ann Arbor, MI

42. Toronto Western Hospital, Toronto, Canada

43. The Ottawa Hospital, Ottawa, Canada

44. Massachusetts General Hospital, Boston, MA

45. University of Kansas Medical Center, Kansas City, KS

46. University of Southern California, Los Angeles, CA

47. Barrow Neurological Institute, Phoenix, AZ

48. Mayo Clinic Arizona, Scottsdale, AZ

49. University of Colorado, Aurora, CO

50. NYU Langone Medical Center, New York, NY

51. University of Florida, Gainesville, FL

52. Montreal Neurological Institute and Hospital/McGill, Montreal, QC, Canada

53. Cleveland Clinic-Las Vegas Lou Ruvo Center for Brain Health, Las Vegas, NV

54. Clinical Ageing Research Unit, Newcastle, UK

55. John Radcliffe Hospital Oxford and Oxford University, Oxford, UK

56. Universität Lübeck, Luebeck, Germany

57. Radboud University, Nijmegen, Netherlands

58. TransThera Consulting

59. Duke University, Durham, NC

60. Wolfson Institute of Population Health, Queen Mary University of London, UK

61. Philipps-University Marburg, Germany

62. University of Lagos, Nigeria

## References

1. Oliveira LMA, Gasser T, Edwards R, et al. Alpha-synuclein research: defining strategic moves in the battle against Parkinson’s disease. NPJ Parkinsons Dis. Jul 26 2021;7(1):65. doi:10.1038/s41531-021-00203-9

2. Pagano G, Ferrara N, Brooks DJ, Pavese N. Age at onset and Parkinson disease phenotype. Neurology. Apr 12 2016;86(15):1400–1407. doi:10.1212/WNL.0000000000002461

3. Postuma RB, Berg D, Stern M, et al. MDS clinical diagnostic criteria for Parkinson’s disease. Mov Disord. Oct 2015;30(12):1591–601. doi:10.1002/mds.26424

4. Gibb WR, Lees AJ. The relevance of the Lewy body to the pathogenesis of idiopathic Parkinson’s disease. J Neurol Neurosurg Psychiatry. Jun 1988;51(6):745–52. doi:10.1136/jnnp.51.6.745

5. Calabresi P, Mechelli A, Natale G, Volpicelli-Daley L, Di Lazzaro G, Ghiglieri V. Alpha-synuclein in Parkinson’s disease and other synucleinopathies: from overt neurodegeneration back to early synaptic dysfunction. Cell Death Dis. Mar 1 2023;14(3):176. doi:10.1038/s41419-023-05672-9

6. Kalia LV, Lang AE. Parkinson’s disease. Lancet. Aug 29 2015;386(9996):896–912. doi:10.1016/S0140-6736(14)61393-3

7. Sieber BA, Landis S, Koroshetz W, et al. Prioritized research recommendations from the National Institute of Neurological Disorders and Stroke Parkinson’s Disease 2014 conference. Ann Neurol. Oct 2014;76(4):469–72. doi:10.1002/ana.24261

8. Hely MA, Morris JG, Reid WG, Trafficante R. Sydney Multicenter Study of Parkinson’s disease: non-L-dopa-responsive problems dominate at 15 years. Mov Disord. Feb 2005;20(2):190–9. doi:10.1002/mds.20324

9. Hely MA, Morris JG, Traficante R, Reid WG, O’Sullivan DJ, Williamson PM. The sydney multicentre study of Parkinson’s disease: progression and mortality at 10 years. J Neurol Neurosurg Psychiatry. Sep 1999;67(3):300–7. doi:10.1136/jnnp.67.3.300

10. Hely MA, Reid WG, Adena MA, Halliday GM, Morris JG. The Sydney multicenter study of Parkinson’s disease: the inevitability of dementia at 20 years. Mov Disord. Apr 30 2008;23(6):837–44. doi:10.1002/mds.21956

11. Gallagher J, Gochanour C, Caspell-Garcia C, et al. Long-Term Dementia Risk in Parkinson Disease. Neurology. Sep 10 2024;103(5):e209699. doi:10.1212/WNL.0000000000209699

12. Siderowf A, Concha-Marambio L, Lafontant DE, et al. Assessment of heterogeneity among participants in the Parkinson’s Progression Markers Initiative cohort using alpha-synuclein seed amplification: a cross-sectional study. Lancet Neurol. May 2023;22(5):407–417. doi:10.1016/S1474-4422(23)00109-6

13. Bellomo G, De Luca CMG, Paoletti FP, Gaetani L, Moda F, Parnetti L. alpha-Synuclein Seed Amplification Assays for Diagnosing Synucleinopathies: The Way Forward. Neurology. Aug 2 2022;99(5):195–205. doi:10.1212/WNL.0000000000200878

14. Brockmann K, Quadalti C, Lerche S, et al. Association between CSF alpha-synuclein seeding activity and genetic status in Parkinson’s disease and dementia with Lewy bodies. Acta Neuropathol Commun. Oct 30 2021;9(1):175. doi:10.1186/s40478-021-01276-6

15. Grossauer A, Hemicker G, Krismer F, et al. alpha-Synuclein Seed Amplification Assays in the Diagnosis of Synucleinopathies Using Cerebrospinal Fluid-A Systematic Review and Meta-Analysis. Mov Disord Clin Pract. May 2023;10(5):737–747. doi:10.1002/mdc3.13710

16. Rossi M, Candelise N, Baiardi S, et al. Ultrasensitive RT-QuIC assay with high sensitivity and specificity for Lewy body-associated synucleinopathies. Acta Neuropathol. Jul 2020;140(1):49–62. doi:10.1007/s00401-020-02160-8

17. Bargar C, Wang W, Gunzler SA, et al. Streamlined alpha-synuclein RT-QuIC assay for various biospecimens in Parkinson’s disease and dementia with Lewy bodies. Acta Neuropathol Commun. Apr 7 2021;9(1):62. doi:10.1186/s40478-021-01175-w

18. Simuni T, Chahine LM, Poston K, et al. A biological definition of neuronal alpha-synuclein disease: towards an integrated staging system for research. Lancet Neurol. Feb 2024;23(2):178–190. doi:10.1016/S1474-4422(23)00405-2

19. Therriault J, Schindler SE, Salvado G, et al. Biomarker-based staging of Alzheimer disease: rationale and clinical applications. Nat Rev Neurol. Apr 2024;20(4):232–244. doi:10.1038/s41582-024-00942-2

20. Therriault J, Zimmer ER, Benedet AL, Pascoal TA, Gauthier S, Rosa-Neto P. Staging of Alzheimer’s disease: past, present, and future perspectives. Trends Mol Med. Sep 2022;28(9):726–741. doi:10.1016/j.molmed.2022.05.008

21. Tabrizi SJ, Schobel S, Gantman EC, et al. A biological classification of Huntington’s disease: the Integrated Staging System. Lancet Neurol. Jul 2022;21(7):632–644. doi:10.1016/S1474-4422(22)00120-X

22. Parkinson Progression Marker I. The Parkinson Progression Marker Initiative (PPMI). Prog Neurobiol. Dec 2011;95(4):629–35. doi:10.1016/j.pneurobio.2011.09.005

23. Brumm MC, Siderowf A, Simuni T, et al. Parkinson’s Progression Markers Initiative: A Milestone-Based Strategy to Monitor Parkinson’s Disease Progression. J Parkinsons Dis. 2023;13(6):899–916. doi:10.3233/JParkinson'sDisease-223433

24. Marek K, Chowdhury S, Siderowf A, et al. The Parkinson’s progression markers initiative (PPMI) - establishing a Parkinson’s Disease biomarker cohort. Ann Clin Transl Neurol. Dec 2018;5(12):1460–1477. doi:10.1002/acn3.644

25. Goetz CG, Tilley BC, Shaftman SR, et al. Movement Disorder Society-sponsored revision of the Unified Parkinson’s Disease Rating Scale (MDS-UPDRS): scale presentation and clinimetric testing results. Mov Disord. Nov 15 2008;23(15):2129–70. doi:10.1002/mds.22340

26. Hoehn MM, Yahr MD. Parkinsonism: onset, progression and mortality. Neurology. May 1967;17(5):427–42. doi:10.1212/wnl.17.5.427

27. Schwab RS EA. Projection technique for evaluating surgery in Parkinson’s disease. 1969:pp.152–157.

28. Doty RLS, P.; Kimmelman, C.P.; Dann, M.S. Olfactory testing as an aid in the diagnosis of Parkinson’s disease: development of optimal discrimination criteria. Neurodegeneration. 1995;4(1):93–7.

29. Visser MM, J.; Stiggelbout, A.M.; van Hilten, J.J. Assessment of Autonomic Dysfunction in Parkinson’s Disease: The SCOPA-AUT. Mov Disord. 2004;19(11):1306–1312.

30. Yesavage JAB, T.L.; Rose, T.L.; Lum, O.; Huang, V.; Adey, M.; Leirer, V.O. Development and validation of a geriatric depression screening scale: a preliminary report. J Psychiatr Res. 1982;17(1):37–49.

31. Stiasny-Kolster KM, G.; Schäfer, S.; Carsten Möller, J.; Heinzel-Gutenbrunner, M.; Oertel, W.H. The REM sleep behavior disorder screening questionnaire--a new diagnostic instrument. Mov Disord. 2007;22(16):2386–93.

32. Jost STK, M.A.; Antonini, A; Martinez-Martin, P. Levodopa Dose Equivalency in Parkinson’s Disease: Updated Systematic Review and Proposals. Mov Disord. 2023;38(7):1236–1252.

33. Nasreddine ZS, Phillips NA, Bedirian V, et al. The Montreal Cognitive Assessment, MoCA: a brief screening tool for mild cognitive impairment. J Am Geriatr Soc. Apr 2005;53(4):695–9. doi:10.1111/j.1532-5415.2005.53221.x

34. Weintraub DC-G, C.; Simuni, T.; Cho, H.R. Neuropsychiatric symptoms and cognitive abilities over the initial quinquennium of Parkinson disease. Ann Clin Transl Neurol. 2020;7(4):449–461.

35. Benedict RHBS, D.; Groninger, L.; Brandt, J. Hopkins Verbal Learning Test—Revised: Normative data and analysis of inter-form and test–retest reliability. Clinical Neuropsychologist. 1998;12(1):43–55.

36. Benton ALH, H.J.; Varney, N.R. Visual perception of line direction in patients with unilateral brain disease. Neurology. 1975;25(10):907–910.

37. Smith A. Symbol Digit Modalities Test: Manual. Western Psychological Services. Los Angeles1982.

38. Wechsler D. Letter-Number Sequencing Subtest. Wechsler Adult Intelligence Scale IV. Pearson; 2008.

39. Benton ALdH, S.K.; Sivan, A.B. Multilingual aphasia examination. 2nd ed. ed. AJA Associates; 1983.

40. Litvan I,; Goldman, J.G.; Tröster, A.I.; Schmand, B.A. Diagnostic criteria for mild cognitive impairment in Parkinson’s disease: Movement Disorder Society Task Force guidelines. Mov Disord. 2012;27(3):349–56.

41. Emre MA, D.; Brown, R.; Burn, D.J. Clinical diagnostic criteria for dementia associated with Parkinson’s disease. 2007;22(12):1689–707.

42. Cousins KAQ, Irwin DJ, Tropea TF, et al. Evaluation of ATN(Parkinson’s Disease) Framework and Biofluid Markers to Predict Cognitive Decline in Early Parkinson Disease. Neurology. Feb 2024;102(4):e208033. doi:10.1212/WNL.0000000000208033

43. Stebbins GT, Goetz CG, Burn DJ, Jankovic J, Khoo TK, Tilley BC. How to identify tremor dominant and postural instability/gait difficulty groups with the movement disorder society unified Parkinson’s disease rating scale: comparison with the unified Parkinson’s disease rating scale. Mov Disord. May 2013;28(5):668–70. doi:10.1002/mds.25383

44. van der Heeden JF, Marinus J, Martinez-Martin P, Rodriguez-Blazquez C, Geraedts VJ, van Hilten JJ. Postural instability and gait are associated with severity and prognosis of Parkinson disease. Neurology. Jun 14 2016;86(24):2243–50. doi:10.1212/WNL.0000000000002768

45. Fereshtehnejad SM, Zeighami Y, Dagher A, Postuma RB. Clinical criteria for subtyping Parkinson’s disease: biomarkers and longitudinal progression. Brain. Jul 1 2017;140(7):1959–1976. doi:10.1093/brain/awx118

46. Macleod AD, Dalen I, Tysnes OB, Larsen JP, Counsell CE. Development and validation of prognostic survival models in newly diagnosed Parkinson’s disease. Mov Disord. Jan 2018;33(1):108–116. doi:10.1002/mds.27177

47. van Rooden SM, Heiser WJ, Kok JN, Verbaan D, van Hilten JJ, Marinus J. The identification of Parkinson’s disease subtypes using cluster analysis: a systematic review. Mov Disord. Jun 15 2010;25(8):969–78. doi:10.1002/mds.23116

48. Lawton M, Ben-Shlomo Y, May MT, et al. Developing and validating Parkinson’s disease subtypes and their motor and cognitive progression. J Neurol Neurosurg Psychiatry. Dec 2018;89(12):1279–1287. doi:10.1136/jnnp-2018-318337

49. Dadu A, Satone V, Kaur R, et al. Identification and prediction of Parkinson’s disease subtypes and progression using machine learning in two cohorts. NPJ Parkinsons Dis. Dec 16 2022;8(1):172. doi:10.1038/s41531-022-00439-z

50. Zhang X, Chou J, Liang J, et al. Data-Driven Subtyping of Parkinson’s Disease Using Longitudinal Clinical Records: A Cohort Study. Sci Rep. Jan 28 2019;9(1):797. doi:10.1038/s41598-018-37545-z

51. Sotirakis C, Su Z, Brzezicki MA, et al. Identification of motor progression in Parkinson’s disease using wearable sensors and machine learning. NPJ Parkinsons Dis. Oct 7 2023;9(1):142. doi:10.1038/s41531-023-00581-2

52. Mestre TA, Fereshtehnejad SM, Berg D, et al. Parkinson’s Disease Subtypes: Critical Appraisal and Recommendations. J Parkinsons Dis. 2021;11(2):395–404. doi:10.3233/JParkinson'sDisease-202472

53. Evans JR, Mason SL, Williams-Gray CH, et al. The natural history of treated Parkinson’s disease in an incident, community based cohort. J Neurol Neurosurg Psychiatry. Oct 2011;82(10):1112–8. doi:10.1136/jnnp.2011.240366

54. Kim HJ, Mason S, Foltynie T, Winder-Rhodes S, Barker RA, Williams-Gray CH. Motor complications in Parkinson’s disease: 13-year follow-up of the CamPaIGN cohort. Mov Disord. Jan 2020;35(1):185–190. doi:10.1002/mds.27882

55. Williams-Gray CH, Evans JR, Goris A, et al. The distinct cognitive syndromes of Parkinson’s disease: 5 year follow-up of the CamPaIGN cohort. Brain. Nov 2009;132(Pt 11):2958–69. doi:10.1093/brain/awp245

56. Caslake R, Taylor K, Scott N, et al. Age-, gender-, and socioeconomic status-specific incidence of Parkinson’s disease and parkinsonism in northeast Scotland: the PINE study. Parkinsonism Relat Disord. May 2013;19(5):515–21. doi:10.1016/j.parkreldis.2013.01.014

57. Counsell C, Giuntoli C, Khan QI, Maple-Grodem J, Macleod AD. The incidence, baseline predictors, and outcomes of dementia in an incident cohort of Parkinson’s disease and controls. J Neurol. Aug 2022;269(8):4288–4298. doi:10.1007/s00415-022-11058-2

58. Williams-Gray CH, Mason SL, Evans JR, et al. The CamPaIGN study of Parkinson’s disease: 10-year outlook in an incident population-based cohort. J Neurol Neurosurg Psychiatry. Nov 2013;84(11):1258–64. doi:10.1136/jnnp-2013-305277

59. Fielding S, Macleod AD, Counsell CE. Medium-term prognosis of an incident cohort of parkinsonian patients compared to controls. Parkinsonism Relat Disord. Nov 2016;32:36–41. doi:10.1016/j.parkreldis.2016.08.010

60. Litvan I, Mangone CA, McKee A, et al. Natural history of progressive supranuclear palsy (Steele-Richardson-Olszewski syndrome) and clinical predictors of survival: a clinicopathological study. J Neurol Neurosurg Psychiatry. Jun 1996;60(6):615–20. doi:10.1136/jnnp.60.6.615

61. Aiba I, Hayashi Y, Shimohata T, et al. Clinical course of pathologically confirmed corticobasal degeneration and corticobasal syndrome. Brain Commun. 2023;5(6):fcad296. doi:10.1093/braincomms/fcad296

62. Picillo M, Barone P, Pellecchia MT, et al. Evolution of mild cognitive impairment in Parkinson disease. Neurology. Apr 15 2014;82(15):1384. doi:10.1212/WNL.0000000000000234

63. Pedersen KF, Larsen JP, Tysnes OB, Alves G. Natural course of mild cognitive impairment in Parkinson disease: A 5-year population-based study. Neurology. Feb 21 2017;88(8):767–774. doi:10.1212/WNL.0000000000003634

64. Jackson H, Anzures-Cabrera J, Taylor KI, Pagano G, Investigators P, Prasinezumab Study G. Hoehn and Yahr Stage and Striatal Dat-SPECT Uptake Are Predictors of Parkinson’s Disease Motor Progression. Front Neurosci. 2021;15:765765. doi:10.3389/fnins.2021.765765

65. Siderowf A, Jennings D, Stern M, et al. Clinical and Imaging Progression in the PARS Cohort: Long-Term Follow-up. Mov Disord. Sep 2020;35(9):1550–1557. doi:10.1002/mds.28139

66. Mollenhauer B, Locascio JJ, Schulz-Schaeffer W, Sixel-Doring F, Trenkwalder C, Schlossmacher MG. alpha-Synuclein and tau concentrations in cerebrospinal fluid of patients presenting with parkinsonism: a cohort study. Lancet Neurol. Mar 2011;10(3):230–40. doi:10.1016/S1474-4422(11)70014-X

67. Kang JH, Irwin DJ, Chen-Plotkin AS, et al. Association of cerebrospinal fluid beta-amyloid 1-42, T-tau, P-tau181, and alpha-synuclein levels with clinical features of drug-naive patients with early Parkinson disease. JAMA Neurol. Oct 2013;70(10):1277–87. doi:10.1001/jamaneurol.2013.3861

68. Hall S, Ohrfelt A, Constantinescu R, et al. Accuracy of a panel of 5 cerebrospinal fluid biomarkers in the differential diagnosis of patients with dementia and/or parkinsonian disorders. Arch Neurol. Nov 2012;69(11):1445–52. doi:10.1001/archneurol.2012.1654

69. Barro C, Chitnis T, Weiner HL. Blood neurofilament light: a critical review of its application to neurologic disease. Ann Clin Transl Neurol. Dec 2020;7(12):2508–2523. doi:10.1002/acn3.51234

70. Backstrom D, Linder J, Jakobson Mo S, et al. NfL as a biomarker for neurodegeneration and survival in Parkinson disease. Neurology. Aug 18 2020;95(7):e827–e838. doi:10.1212/WNL.0000000000010084

71. Mollenhauer B, Dakna M, Kruse N, et al. Validation of Serum Neurofilament Light Chain as a Biomarker of Parkinson’s Disease Progression. Mov Disord. Nov 2020;35(11):1999–2008. doi:10.1002/mds.28206

72. Frank A, Bendig J, Schnalke N, et al. Serum neurofilament indicates accelerated neurodegeneration and predicts mortality in late-stage Parkinson’s disease. NPJ Parkinsons Dis. Jan 9 2024;10(1):14. doi:10.1038/s41531-023-00605-x

73. Pagano G, Taylor KI, Anzures-Cabrera J, et al. Trial of Prasinezumab in Early-Stage Parkinson’s Disease. N Engl J Med. Aug 4 2022;387(5):421–432. doi:10.1056/NEJMoa2202867

74. Lang AE, Siderowf AD, Macklin EA, et al. Trial of Cinpanemab in Early Parkinson’s Disease. N Engl J Med. Aug 4 2022;387(5):408–420. doi:10.1056/NEJMoa2203395

